# Genetic Determinants of Pulmonary Artery Size in over 50,000 Subjects with and without COPD

**DOI:** 10.64898/2026.07.01.26357039

**Authors:** Vasile Foris, Kangjin Kim, Courtney Tern, Yunzhe Qian, Junsun Yu, George Washko, R Chad Wade, J. Michael Wells, Honghuang Lin, George T. O’Connor, Albert V. Smith, Stacey B. Gabriel, Namrata Gupta, Edwin K. Silverman, Adel Boueiz, Michael H. Cho

## Abstract

**Rationale:** Pulmonary artery (PA) enlargement is a non-invasive imaging biomarker associated with pulmonary hypertension and mortality in COPD; however, its genetic determinants remain incompletely understood.

**Objectives:** To characterize the genetic architecture of PA size across COPD-enriched and population-based cohorts.

**Methods:** We performed genome-wide association analyses of PA diameter using whole-genome sequencing in COPDGene (n=9,418) and ECLIPSE (n=1,859), and imputed-genotype data from the UK Biobank (n=37,073). We replicated lead variants in the Framingham Heart Study (FHS; n=3,289), incorporated all four studies into a joint meta-analysis, and identified independent signals through conditional analyses. Candidate effector genes were prioritized using coding variant annotation, colocalization, and integrative regulatory evidence.

**Measurements and Main Results:** We identified 44 independent genome-wide significant PA diameter signals within 39 loci, including 8 variants replicated in FHS, novel associations near *FRMD4B*, *SLC20A2*, *BORCS7-ASMT*, and *KCNRG*, and 5 signals in conditional analysis including multiple signals at *ANO1*. Genetic effects were concordant across imaging modalities and cohorts of differing COPD burden. Effector-gene prioritization nominated *ABCC8*, *PDGFD*, *HMCN1*, *CCNE1*, and *TBX20*, implicating pathways in vascular remodeling, developmental regulation, smooth muscle and endothelial function, ion-channel signaling, and extracellular matrix organization. Colocalization with pulse pressure GWAS demonstrated substantial shared causal variation between pulmonary and systemic vascular biology.

**Conclusions:** In this largest genetic study of pulmonary vascular imaging to date, PA diameter exhibits a polygenic architecture consistent across imaging modalities and cohorts of differing COPD burden. The prioritized effector genes bridge rare-variant pulmonary hypertension biology with common-variant systemic vascular biology.

## INTRODUCTION

Pulmonary artery (PA) size on chest CT is an established marker of pulmonary vascular disease. Measured by PA diameter or PA-to-aorta ratio (PA/A), it robustly predicts pulmonary hypertension (PH), respiratory exacerbations, right heart dysfunction, and mortality across diverse populations (1–3). In smokers and individuals with chronic obstructive pulmonary disease (COPD), PA enlargement is a non-invasive indicator of pulmonary vascular involvement and adverse cardiopulmonary outcomes (4–12). These features position PA size as a scalable and clinically useful trait for risk stratification and pulmonary vascular disease characterization.

Despite its clinical relevance, the genetic architecture of PA size remains incompletely defined, particularly in COPD. While both common and rare genetic variants are established contributors to PH susceptibility (13), the genetic determinants of PA size itself have been minimally investigated. A prior genome-wide association study (GWAS) identified common variants near *IREB2* and *GALC* associated with PA enlargement in COPD (14). More recent GWAS of MRI-derived right heart phenotypes in the UK Biobank identified additional loci associated with PA traits as part of broader cardiac chamber analyses (15–18). However, these efforts did not specifically interrogate smokers or individuals with COPD, and provided limited insight into causal variants, effector genes, biological pathways, or tissue-specific mechanisms.

We hypothesized that common and rare genetic variation influencing PA size converges on biological pathways linking pulmonary vascular remodeling, systemic vascular biology, and cardiopulmonary structure across both COPD-enriched and general populations. The availability of deeply phenotyped COPD-enriched cohorts, together with advances in statistical fine-mapping and functional genomic annotation, now enables a more comprehensive characterization of PA size genetics. We conducted a large-scale genetic study of PA size in over 50,000 individuals with and without COPD, and integrated multi-omic functional data to prioritize causal variants, effector genes, and tissue contexts underlying PA remodeling. Some of these results have been previously reported in abstract form (19).

## MATERIALS AND METHODS

### Cohorts and phenotype definitions

COPDGene and ECLIPSE are COPD-enriched cohorts of current and former smokers. Maximal main PA diameter and PA-to-aorta (PA/A) ratio were measured on chest CT scans at the level of the PA bifurcation (11,20,21). Whole genome sequencing were obtained through the NHLBI Trans-Omics in Precision Medicine (TOPMed) (22). The UK Biobank systolic PA diameter derived from deep learning-based segmentation of cardiac MRI GWAS summary statistics were obtained from the Cardiovascular Disease Knowledge Portal (15,23,24). We additionally used GWAS summary statistics for CT measured maximum main PA diameter from the Framingham Heart Study (FHS) (25). PA diameter was selected as the primary trait because it was available across all cohorts and represented the most comparable phenotype across imaging modalities. All studies received institutional review board approval, and all participants provided written informed consent. Additional cohort details are provided in the Online Supplement.

### Association testing

We employed a three-stage discovery-replication design followed by a locus interpretation framework depicted in **Figure 1**. In Stage 1, common- and rare-variant association analyses were performed in the combined COPDGene and ECLIPSE cohort using REGENIE v4 (26). Models were adjusted for age, sex, height, study cohort, sequencing center, smoking pack-years, FEV₁, body mass index, and the first 20 genetic ancestry principal components, with mean-centered squared terms for age and height. Genome-wide significance was defined as P<5×10⁻⁸ for single-variant analyses for common variants (minor allele frequency (MAF)>0.01) and P<2.5×10⁻⁶ for gene-based rare-variant analyses. Secondary analyses of PA/A and PA-enlargement, as well as cohort-specific, COPD-stratified analyses and rare-variant analyses are described in the Online Supplement.

**Figure 1.**
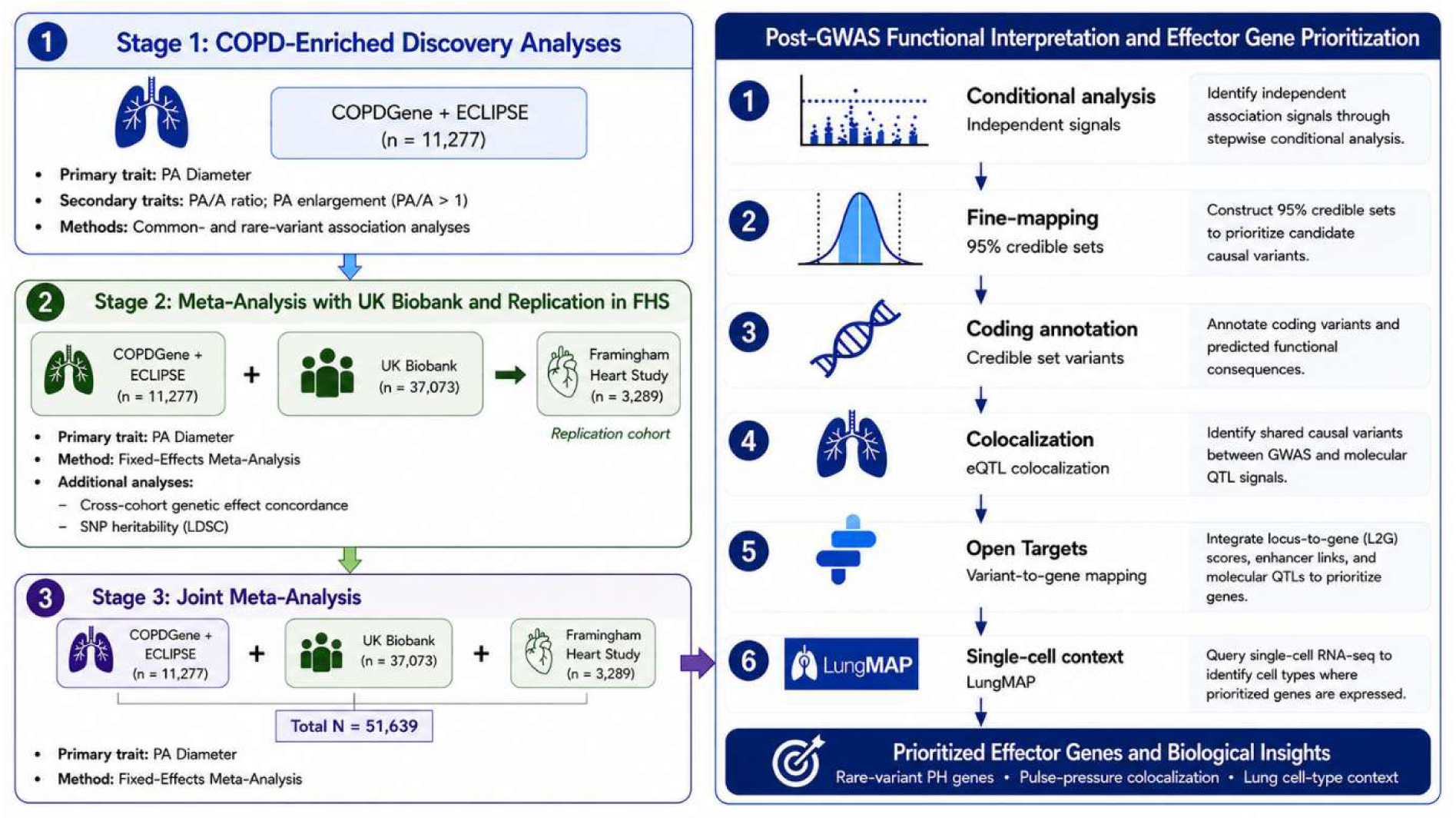
Study design and post-GWAS locus interpretation framework. Overview of the multi-cohort analytical framework used to identify genetic loci associated with pulmonary artery (PA) diameter and prioritize candidate effector genes. Stage 1 comprised discovery analyses in the COPDGene and ECLIPSE cohorts. Stage 2 included meta-analysis with the UK Biobank and independent replication in the FHS Framingham Heart Study. In Stage 3, a joint fixed-effects meta-analysis across all cohorts was performed. Post-GWAS analyses included conditional analysis to identify independent association signals, Bayesian fine-mapping, coding-variant annotation, eQTL colocalization, Open Targets locus-to-gene prioritization, and single-cell transcriptomic interrogation using LungMAP Single Cell Reference to prioritize biologically supported effector genes and relevant cell types.

### Meta-analyses, replication, and heritability estimation

In Stage 2, fixed-effects inverse-variance weighted meta-analysis of COPDGene, ECLIPSE, and UK Biobank was performed using METAL (27). Cross-modality concordance between CT-derived PA traits (COPDGene/ECLIPSE) and MRI-derived PA traits (UK Biobank) was evaluated using Pearson correlation of effect estimates for variants reaching genome-wide significance in either or both datasets.

Genome-wide significant Stage 2 PA diameter variants were tested for replication in the FHS cohort. Given the smaller sample size of FHS, nominal significance *(one-sided P<0.05)* with a concordant effect direction was considered supportive evidence of replication.

SNP heritability (h^2^) and cross-cohort genetic correlation (rg) were estimated by (linkage disequilibrium) LD score regression, applied separately to the COPD-enriched discovery dataset (COPDGene+ECLIPSE) and the UK Biobank (28,29).

In Stage 3, FHS was incorporated into a joint four-cohort meta-analysis of PA diameter. As downstream analyses including conditional analyses, fine-mapping, and colocalization require ancestry-matched LD structure, we restricted these analyses to participants of self-reported European ancestry using an in-sample LD reference panel derived from COPDGene and ECLIPSE.

### Conditional analysis, fine-mapping, colocalization, and effector gene prioritization

Conditionally independent PA association signals were identified using conditional and joint analysis (GCTA-COJO; LD threshold r² < 0.9) (30,31). Statistical fine-mapping was performed using SuSiE-RSS (32,33). Variants within credible sets were annotated for predicted functional consequence. Bayesian colocalization (coloc.abf; PP.H4≥0.8) was performed between PA diameter signals and GTEx v8 expression quantitative trait loci (eQTL) across cardiopulmonary-relevant tissues (lung, coronary artery, tibial artery, aorta, heart atrial appendage, left ventricle and blood), supplemented by lung eQTL data from the Lung Tissue Research Consortium (34). We additionally performed colocalization analyses with GWAS summary statistics for pulse pressure from the Million Veteran Program and pulmonary arterial hypertension to identify shared vascular genetic effects (35,36). Associations between lead variants and plasma protein QTL (pQTLs) were queried in the UK Biobank Pharma Proteomics Project (37).

For additional evidence linking candidate effector genes and regulatory mechanisms, we queried the Open Targets Genetics (OT) platform. For each conditionally independent lead variant, we extracted credible-set membership with posterior probabilities of causality (PPC), locus-to-gene (L2G) scores, enhancer-to-gene (E2G) scores, and molecular QTL associations spanning expression, protein, splicing (sQTL), single-cell expression (scQTL), and transcript-usage (tuQTL) datasets (38). To assess pleiotropic overlaps across cardiopulmonary and systemic traits OT phenotype annotations were harmonized into biologically coherent categories.

For each signal we then integrated five complementary sources of effector-gene evidence: (1) a qualifying coding variant (missense or nonsense) within the SuSiE-RSS 95% credible set; (2) eQTL colocalization; (3) L2G; (4) E2G; and (5) molecular QTL. Evidence sources contributed only if they met a prespecified high-confidence threshold and nominated a protein-coding gene (GENCODE v38) (39): 1) a qualifying coding variant within the lead’s 95% credible set (missense or nonsense; PIP ≥ 0.10), 2) eQTL colocalization (GTEx v8; PP.H4 ≥ 0.80), OT 3) locus-to-gene (L2G ≥ 0.50), 4) enhancer-to-gene (E2G ≥ 0.60), and 5) molecular QTL evidence (any modality: eQTL, pQTL, sQTL, sc-eQTL, tuQTL). The prioritized effector gene was nominated by the highest-ranking qualifying evidence source, using the predefined hierarchy coding variant >eQTL colocalization > L2G > E2G > molecular QTL, or the nearest protein-coding gene when no evidence source qualified. The converging-layers count (maximum five) was defined as the number of qualifying evidence sources supporting the prioritized gene. Cell-type expression of prioritized genes was assessed using the LungMAP Human Lung CellRef single-cell reference atlas (40). Additional methodological details are provided in the Online Supplement.

## RESULTS

### Study Design and Cohort Characteristics

Our overall study design is shown in **Figure 1** and baseline cohort characteristics in **Table 1**. Stage 1 discovery comprised 9,418 smokers from COPDGene and 1,859 from ECLIPSE (COPD prevalence 45% and 100%, respectively). Stage 2 added 37,073 UK Biobank participants, and Stage 3 added 3,289 FHS participants for replication and joint meta-analysis. Cohort flow diagrams and additional participant characteristics are presented in the Online Supplement (Figure S1, Table S1-2).

**Table 1.**
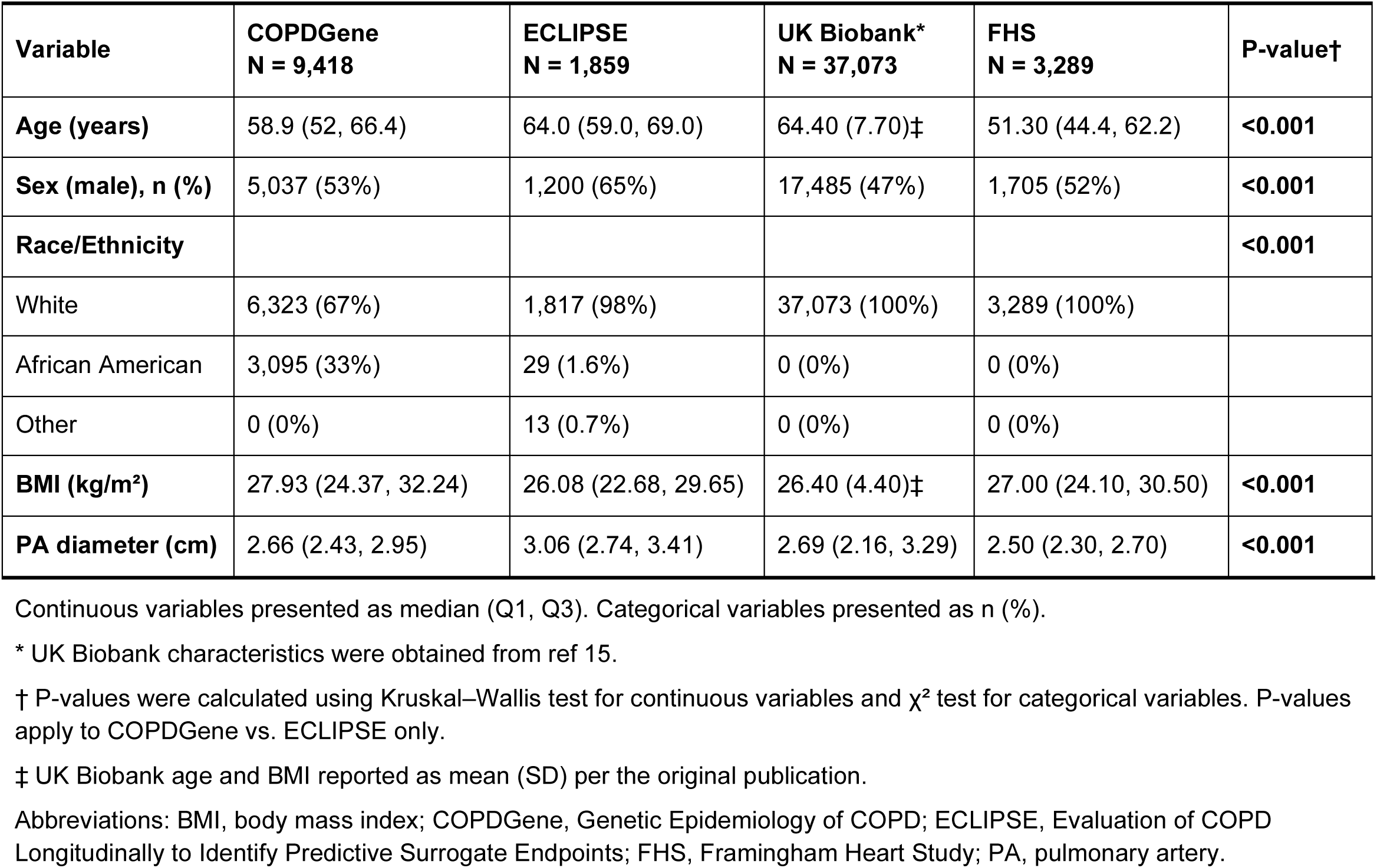
Baseline characteristics of the four study cohorts.

### Genetic Discovery in COPD-Enriched Cohorts and Multi-Cohort Meta-analysis Identify Genome-wide Significant Loci

Genome-wide association analysis in the combined COPDGene and ECLIPSE discovery cohort identified a genome-wide significant locus associated with PA diameter on chromosome 11, with lead variant rs386829462 near *ANO1* (P=1.89×10⁻⁸; Figure S2). For PA/A, one locus on chromosome 2 also surpassed the genome-wide threshold, represented by rs1363065 near *ACYP2* (*P=2.4×10⁻⁸*; Figure S2C). No genome-wide significant associations were observed for the binary PA-enlargement phenotype. The *ANO1* signal was also observed in an analysis of the same PA diameter phenotype from COPDGene Phase 2 (n = 4,848), where the lead variant was in strong LD [r²=0.99] with rs386829462, P=6.87×10⁻⁹; Figure S2E). COPD-stratified and single-cohort analyses revealed no additional loci. Gene-based rare-variant burden testing nominated *GEM* at exome-wide significance in COPDGene (Figure S3, Table S3), whereas no additional exome-wide signals were detected in the combined cohort.

Fixed-effects meta-analysis across COPDGene, ECLIPSE, and the UK Biobank (n=48,350) identified 37 genome-wide significant PA diameter loci (Figure S4A-B, Table S4). Of these 31 mapped to regions previously associated with PA size or related right-heart phenotypes, including the *ANO1* locus identified in Stage 1. The remaining six novel loci were located near *FRMD4B, TSC22D2, SLC20A2, BORCS7-ASMT, KCNRG*, and *TCF12*. Meta-analysis results of PA/A are reported in the Online Supplement (Figure S4C–D, Table S5). Notably, *TCF12* emerged as a shared novel locus across both PA traits, implicating common genetic determinants of pulmonary vascular structure across distinct imaging phenotypes.

To evaluate consistency of genetic associations between CT-derived PA traits in COPD-enriched cohorts and MRI-derived measures in UK Biobank, we compared effect estimates for variants reaching genome-wide significance in either or both datasets. Effect sizes for both PA diameter (Pearson r = 0.89, P = 2.2×10⁻¹⁶; n = 1,132 SNPs; **Figure 2**) and PA/A (r = 0.89, *P = 2.2×10*⁻¹⁶; n = 516 SNPs, Figure S5) showed strong correlation, consistent with reproducible signals across imaging modalities and study populations.

**Figure 2.**
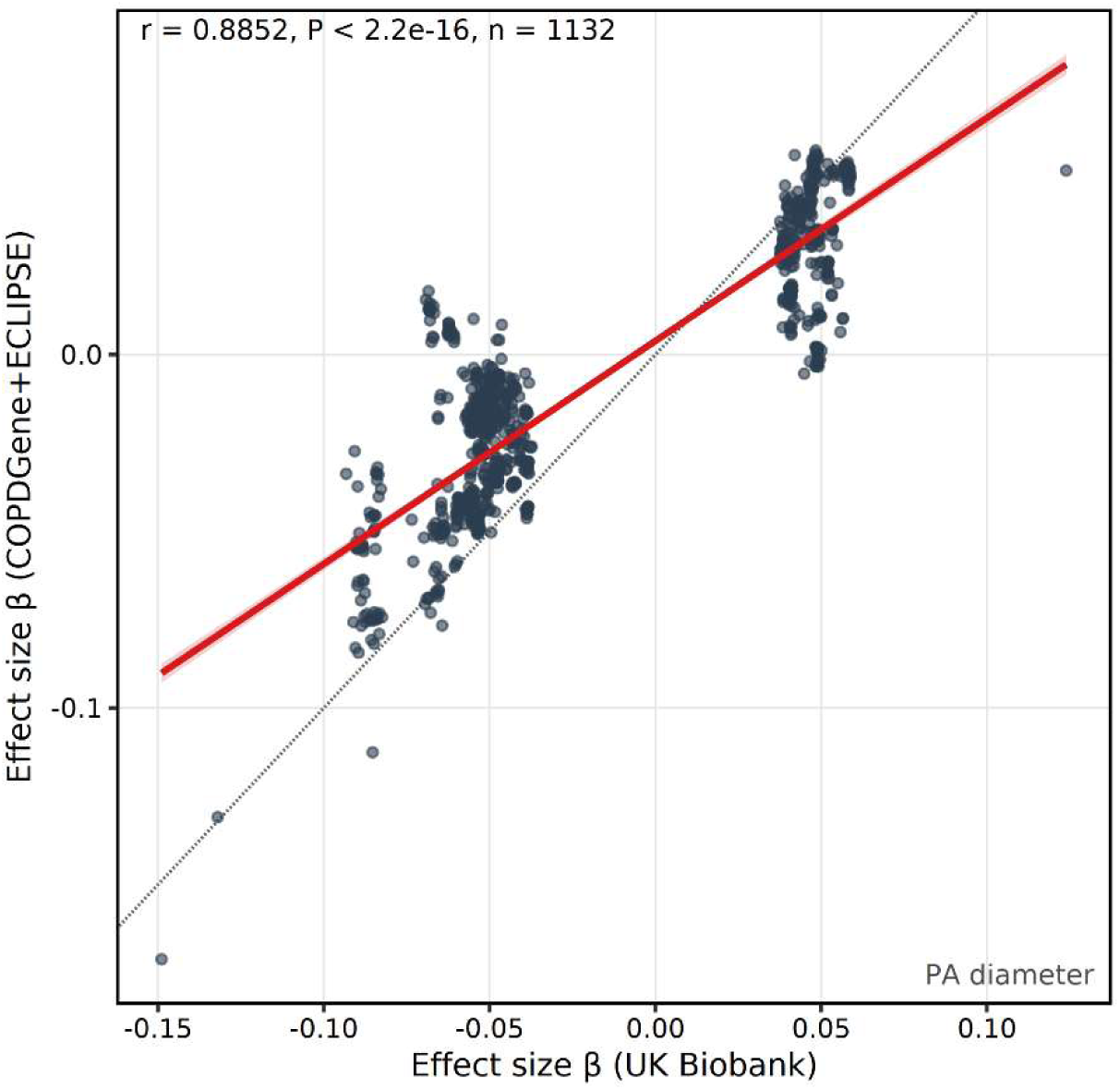
Cross-cohort concordance of genetic effects for pulmonary artery diameter. Cross-cohort correlation of genetic effect estimates for PA diameter between the UK Biobank (x-axis) and combined COPDGene+ECLIPSE discovery cohorts (y-axis) for variants reaching genome-wide significance in either or both dataset. The red line represents the linear regression fit with 95% confidence interval, and the dashed gray line indicates perfect concordance (y = x). Pearson correlation coefficients, P-values, and variant counts are annotated.

### PA Traits Demonstrate Heritability and Shared Genetic Architecture Across Diverse Cohorts

Observed-scale SNP heritability for PA diameter was substantial in the UK Biobank (h²=0.286, SE=0.022) and lower and less precise in the COPD-enriched cohorts (h²=0.107, SE=0.036), likely reflecting smaller sample size and greater environmental and disease-related heterogeneity in smoking-exposed populations. LD score regression intercepts were near 1.0, indicating minimal inflation from population stratification.

Genetic correlation between COPD-enriched and population-based cohorts for PA diameter, was 1 (rg=1.02, SE=0.17, P=3.0×10⁻⁹), indicating highly similar genetic effects despite major differences in smoking exposure and COPD burden. Together, these findings indicate that PA diameter is heritable and largely governed by shared genetic determinants across populations. Heritability and genetic correlation for the secondary PA/A ratio are reported in the Online Supplement (Table S6).

### Independent Replication Supports PA Diameter Associations in the Framingham Heart Study

We sought independent replication in the FHS (n=3,289) for the 37 lead PA diameter variants identified in the Stage 2 discovery meta-analysis. Of these, 36 were available for testing in FHS. Applying the prespecified replication criteria, eight variants met the threshold for nominal replication in FHS *(P< 0.05)*, including four of the six novel loci (*FRMD4B, SLC20A2, BORCS7-ASMT, KCNRG)*. Per-locus replication results are summarized in **Table 2** and Table S7.

**Table 2.** Genomic risk loci for pulmonary artery diameter from the joint meta-analysis, with conditionally independent signals and replication in the Framingham Heart Study.

|  |  |  |  | Joint meta-analysis |  |  |  | FHS replication (Stage 2) |  |  |  |
| --- | --- | --- | --- | --- | --- | --- | --- | --- | --- | --- | --- |
| Locus | Lead variant (GRCh38) | EA/OA | Nearest gene | EAF | $\beta$ | SE | P | FHS $\beta$ | FHS SE | FHS P | Replication (FHS) |
| 1 | 1:59408410:T:C | T/C | *FGGY* | 0.695 | -0.0395 | 0.0064 | $5.59 \times 10^{-10}$ | -0.098 | 0.078 | $2.10 \times 10^{-1}$ | No |
| 2 | 1:185795687:G:A | A/G | *HMCN1* | 0.569 | 0.0432 | 0.0061 | $1.24 \times 10^{-12}$ | 0.187 | 0.072 | $9.40 \times 10^{-3}$ | Yes |
| 3 | 3:41712930:A:T | A/T | *ULK4* | 0.834 | -0.0454 | 0.0078 | $4.80 \times 10^{-9}$ | -0.088 | 0.092 | $3.40 \times 10^{-1}$ | No |
| 4 | 3:69353772:T:G | T/G | *FRMD4B* | 0.692 | 0.0424 | 0.0064 | $2.79 \times 10^{-11}$ | 0.210 | 0.078 | $7.20 \times 10^{-3}$ | Yes |
| 5 | 3:128484100:G:C | C/G | *GATA2* | 0.029 | -0.1083 | 0.0184 | $3.77 \times 10^{-9}$ | -0.235 | 0.232 | $3.10 \times 10^{-1}$ | No |
| Locus | Lead variant (GRCh38) | EA/OA | Nearest gene | EAF | $\beta$ | SE | P | FHS $\beta$ | FHS SE | FHS P | Replication (FHS) |
| 6 | 3:150392939:A:T | A/T | *TSC22D2* | 0.974 | 0.1024 | 0.0166 | $7.45 \times 10^{-10}$ | 0.280 | 0.274 | $3.10 \times 10^{-1}$ | No |
| 7 | 3:169428709:C:T | T/C | *MECOM* | 0.514 | -0.0449 | 0.0059 | $2.86 \times 10^{-14}$ | -0.104 | 0.072 | $1.50 \times 10^{-1}$ | No |
| 8 | 5:96271283:A:C | A/C | *PCSK1* | 0.656 | 0.0344 | 0.0063 | $4.22 \times 10^{-8}$ | -0.021 | 0.075 | $7.80 \times 10^{-1}$ | No |
| 9 | 5:123167178:T:C | T/C | *PRDM6* | 0.821 | 0.0470 | 0.0078 | $1.88 \times 10^{-9}$ | -0.246 | 0.090 | $6.20 \times 10^{-3}$ | Yes † |
| 10 | 5:158834155:C:T | T/C | *EBF1* | 0.513 | -0.0371 | 0.0060 | $5.05 \times 10^{-10}$ | -0.087 | 0.072 | $2.30 \times 10^{-1}$ | No |
| 11 | 5:173235021:T:C | T/C | *NKX2-5* | 0.705 | -0.0427 | 0.0064 | $3.59 \times 10^{-11}$ | -0.051 | 0.085 | $5.50 \times 10^{-1}$ | No |
| 12 | 6:36679072:T:C | T/C | *CDKN1A* | 0.677 | -0.0424 | 0.0063 | $2.56 \times 10^{-11}$ | -0.130 | 0.077 | $8.90 \times 10^{-2}$ | No |
| 13 | 6:143290334:G:T | T/G | *AIG1* | 0.393 | 0.0417 | 0.0063 | $2.55 \times 10^{-11}$ | 0.029 | 0.074 | $6.90 \times 10^{-1}$ | No |
| 14 | 7:35243841:G:C | C/G | *TBX20* | 0.391 | -0.0575 | 0.0060 | $1.53 \times 10^{-21}$ | -0.139 | 0.074 | $6.10 \times 10^{-2}$ | No |
| | 7:35466100:C:G * | C/G | *HERPUD2* | 0.980 | -0.1435 | 0.0206 | $3.53 \times 10^{-12}$ | — | — | — | Not tested |
| 15 | 7:74015790:A:T | A/T | *ELN* | 0.545 | -0.0413 | 0.0060 | $5.63 \times 10^{-12}$ | 0.101 | 0.073 | $1.70 \times 10^{-1}$ | No |
| 16 | 7:85415802:AT:A | A/AT | *SEMA3D* | 0.429 | 0.0373 | 0.0062 | $1.51 \times 10^{-9}$ | 0.015 | 0.073 | $8.30 \times 10^{-1}$ | No |
| 17 | 8:42467247:T:C | T/C | *SLC20A2* | 0.547 | 0.0340 | 0.0059 | $9.40 \times 10^{-9}$ | 0.194 | 0.075 | $9.40 \times 10^{-3}$ | Yes |
| 18 | 8:140049929:G:A | A/G | *TRAPPC9* | 0.367 | 0.0385 | 0.0061 | $2.77 \times 10^{-10}$ | 0.107 | 0.074 | $1.50 \times 10^{-1}$ | No |
| 19 | 10:77418286:G:C | C/G | *KCNMA1* | 0.453 | -0.0334 | 0.0060 | $2.44 \times 10^{-8}$ | -0.087 | 0.073 | $2.30 \times 10^{-1}$ | No |
| 20 | 10:94279840:G:C | C/G | *PLCE1* | 0.447 | 0.0380 | 0.0059 | $1.43 \times 10^{-10}$ | 0.075 | 0.073 | $3.00 \times 10^{-1}$ | No |
| | 10:94311455:T:C * | T/C | *PLCE1* | 0.808 | 0.0603 | 0.0092 | $6.88 \times 10^{-11}$ | — | — | — | Not tested |
| 21 | 10:102903553:G:A | A/G | *BORCS7-ASMT* | 0.308 | 0.0390 | 0.0064 | $1.32 \times 10^{-9}$ | 0.193 | 0.079 | $1.40 \times 10^{-2}$ | Yes |
| 22 | 11:17476510:G:A | A/G | *ABCC8* | 0.181 | 0.0709 | 0.0083 | $1.56 \times 10^{-17}$ | 0.292 | 0.095 | $2.10 \times 10^{-3}$ | Yes |
| 23 | 11:47348490:T:C | T/C | *MYBPC3* | 0.872 | -0.0547 | 0.0089 | $6.86 \times 10^{-10}$ | -0.157 | 0.108 | $1.40 \times 10^{-1}$ | No |
| 24 | 11:70163614:G:A | A/G | *ANO1* | 0.408 | -0.0418 | 0.0061 | $9.22 \times 10^{-12}$ | -0.105 | 0.073 | $1.50 \times 10^{-1}$ | No |
| | 11:69960164:G:A * | A/G | *ANO1* | 0.160 | 0.0463 | 0.0082 | $2.00 \times 10^{-8}$ | — | — | — | Not tested |
| | 11:69976005:G:T * | T/G | *ANO1* | 0.081 | 0.0642 | 0.0113 | $1.18 \times 10^{-8}$ | — | — | — | Not tested |
| 25 | 11:103789839:T:C | T/C | *PDGFD* | 0.705 | 0.0479 | 0.0064 | $8.65 \times 10^{-14}$ | 0.071 | 0.078 | $3.60 \times 10^{-1}$ | No |
| 26 | 12:20077705:A:G | A/G | *PDE3A* | 0.801 | -0.0464 | 0.0076 | $8.30 \times 10^{-10}$ | -0.278 | 0.093 | $2.90 \times 10^{-3}$ | Yes |
| 27 | 12:21855433:G:T | T/G | *ABCC9* | 0.410 | -0.0338 | 0.0061 | $3.62 \times 10^{-8}$ | — | — | — | Not tested |
| 28 | 12:50919064:T:C | T/C | *METTL7A* | 0.654 | -0.0344 | 0.0063 | $4.01 \times 10^{-8}$ | — | — | — | Not tested |
| 29 | 12:66000884:C:T | T/C | *HMGA2* | 0.380 | 0.0455 | 0.0060 | $5.01 \times 10^{-14}$ | 0.009 | 0.074 | $9.10 \times 10^{-1}$ | No |
| | 12:65858514:A:G * | A/G | *HMGA2* | 0.863 | -0.0542 | 0.0089 | $1.17 \times 10^{-9}$ | — | — | — | Not tested |
| 30 | 12:70720620:T:C | T/C | *PTPRR* | 0.461 | -0.0369 | 0.0060 | $6.45 \times 10^{-10}$ | -0.033 | 0.071 | $6.40 \times 10^{-1}$ | No |
| Locus | Lead variant (GRCh38) | EA/OA | Nearest gene | EAF | $\beta$ | SE | P | FHS $\beta$ | FHS SE | FHS P | Replication (FHS) |
| 31 | 12:93769687:C:G | C/G | *CRADD* | 0.566 | 0.0380 | 0.0060 | $2.76 \times 10^{-10}$ | 0.059 | 0.073 | $4.20 \times 10^{-1}$ | No |
| 32 | 12:111427245:C:G | C/G | *SH2B3* | 0.502 | -0.0389 | 0.0061 | $1.91 \times 10^{-10}$ | 0.136 | 0.072 | $6.00 \times 10^{-2}$ | No |
| 33 | 13:50199654:C:T | T/C | *KCNRG* | 0.532 | 0.0414 | 0.0059 | $1.98 \times 10^{-12}$ | 0.170 | 0.072 | $1.80 \times 10^{-2}$ | Yes |
| 34 | 15:56925420:A:G | A/G | *TCF12* | 0.469 | 0.0351 | 0.0061 | $7.85 \times 10^{-9}$ | 0.071 | 0.073 | $3.30 \times 10^{-1}$ | No |
| 35 | 17:2125634:A:G | A/G | *SMG6* | 0.826 | -0.0667 | 0.0078 | $7.78 \times 10^{-18}$ | -0.108 | 0.099 | $2.70 \times 10^{-1}$ | No |
| 36 | 19:11165166:G:A | A/G | *KANK2* | 0.325 | 0.0422 | 0.0065 | $6.39 \times 10^{-11}$ | 0.080 | 0.078 | $3.00 \times 10^{-1}$ | No |
| 37 | 19:29822669:C:T | T/C | *CCNE1* | 0.392 | 0.0363 | 0.0061 | $3.46 \times 10^{-9}$ | 0.099 | 0.075 | $1.90 \times 10^{-1}$ | No |
| 38 | 19:38659595:A:G | A/G | *ACTN4* | 0.546 | -0.0432 | 0.0060 | $5.23 \times 10^{-13}$ | 0.019 | 0.073 | $8.00 \times 10^{-1}$ | No |
| 39 | 22:38181508:G:A | A/G | *PLA2G6* | 0.439 | -0.0344 | 0.0062 | $2.91 \times 10^{-8}$ | — | — | — | Not tested |

Each primary row is one genomic risk locus ( $n = 39$ ) from the joint meta-analysis of COPDGene, ECLIPSE, UK Biobank and FHS; $\beta$ , SE and P are joint-meta estimates.
\* Additional conditionally independent signal at the locus from GCTA-COJO conditional analysis in non-Hispanic White participants; $\beta$ , SE and P are conditional estimates. These signals were not individually tested in FHS.
† At the PRDM6 locus, the FHS estimate shown is for 5:123141240:C:A (the Stage 2 lead, not the joint-meta lead): $\beta = -0.246$ , SE = 0.090, P = $6.20 \times 10^{-3}$ .
“Not tested” indicates the variant was not assessed in FHS, because the locus was not genome-wide significant in the Stage 2 (FHS-excluded) meta-analysis or because the row is a secondary conditional signal. *PLA2G6* reached genome-wide significance only in European ancestry analysis; its estimates are specific to that analysis.
Dash (—) indicates not applicable.
EAF is the effect allele frequency in the European ancestry COPDGene and ECLIPSE reference panel.
Definition of abbreviations: $\beta$ = per-allele effect size; COJO = conditional and joint analysis; EA = effect allele; EAF = effect allele frequency; FHS = Framingham Heart Study; OA = other allele; P = P-value; SE = standard error.

### Joint Meta-analysis and Conditional Analysis Identify Additional PA Loci and Allelic Heterogeneity

The four-cohort joint meta-analysis (n=51,639) identified 38 genome-wide significant PA diameter loci (**Figure 3**; QQ plot in Figure S6A), adding a locus near *METTL7A* relative to the Stage 2 meta-analysis. Restricting to participants of European ancestry for the LD-dependent downstream analyses yielded one additional locus near *PLA2G6*, while three loci from the multi-ancestry analysis (*METTL7A, ULK4, KCNMA1*) were no longer significant. In total, 39 loci were associated with PA diameter across the multi-ancestry and European-ancestry analyses (Figure S6B-C). Conditional analyses using GCTA-COJO further refined this architecture, identifying 41 conditionally independent signals across the 36 European loci, including 5 secondary signals at four loci. Notably, *ANO1* harbored three independent signals, while *PLCE1, HMGA2, and TBX20/HERPUD2* each contained two (**Table 2**; Table S8). Thus, combining independent signals identified in the multi-ancestry or European ancestry analyses, we identified 44 independent genome-wide significant associations across the 39 loci, highlighting the highly polygenic and genetically complex architecture of pulmonary artery structure.

**Figure 3.**
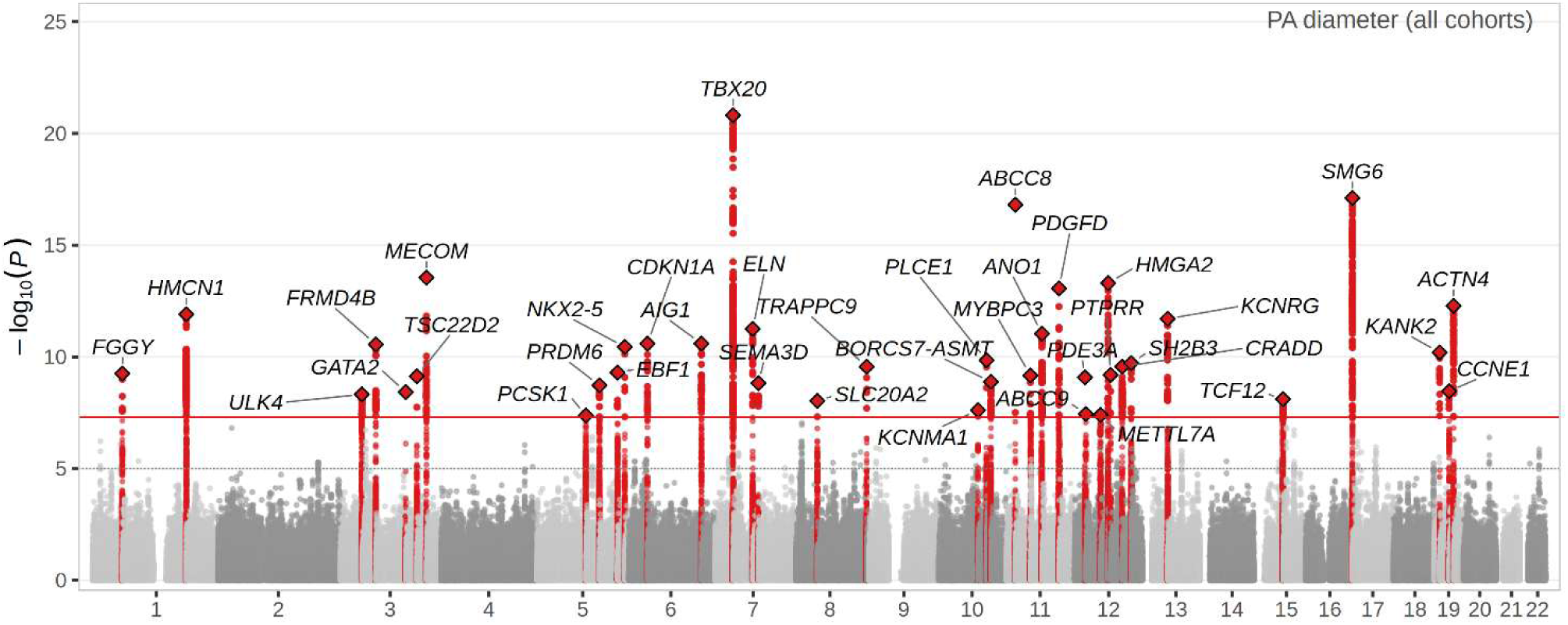
Stage 3 joint meta-analysis of PA diameter across all four cohorts. Manhattan plot of the fixed-effects, inverse-variance weighted meta-analysis for PA diameter combining COPDGene, ECLIPSE, the UK Biobank, and the Framingham Heart Study (total n = 51,639). Variants within genome-wide significant loci (±500 kb of the lead variant) are highlighted in red. Lead variants are shown as diamonds and annotated by the nearest protein-coding gene. The solid red line denotes genome-wide significance (P = 5×10⁻⁸), and the dashed gray line indicates the suggestive significance threshold (P = 1×10⁻⁵).

### Statistical Fine-Mapping Refines Candidate Causal Variants at PA Diameter Loci

As many variant-to-gene approaches are ancestry specific, subsequent analyses were restricted to the 41 independent loci identified in European ancestry subjects. We integrated five complementary sources of evidence for effector-gene prioritization (see Methods).

First, to refine association signals and prioritize likely causal variants, we performed Bayesian fine-mapping using SuSiE-RSS. Fine-mapping identified 45 credible sets, with a median size of 8 variants. Seven loci contained a lead variant with a posterior inclusion probability (PIP) ≥0.5, including 4 loci near *MECOM, PDE3A, and ABCC8* with PIP>0.95 (**Figure 4**; Table S9).

**Figure 4.**
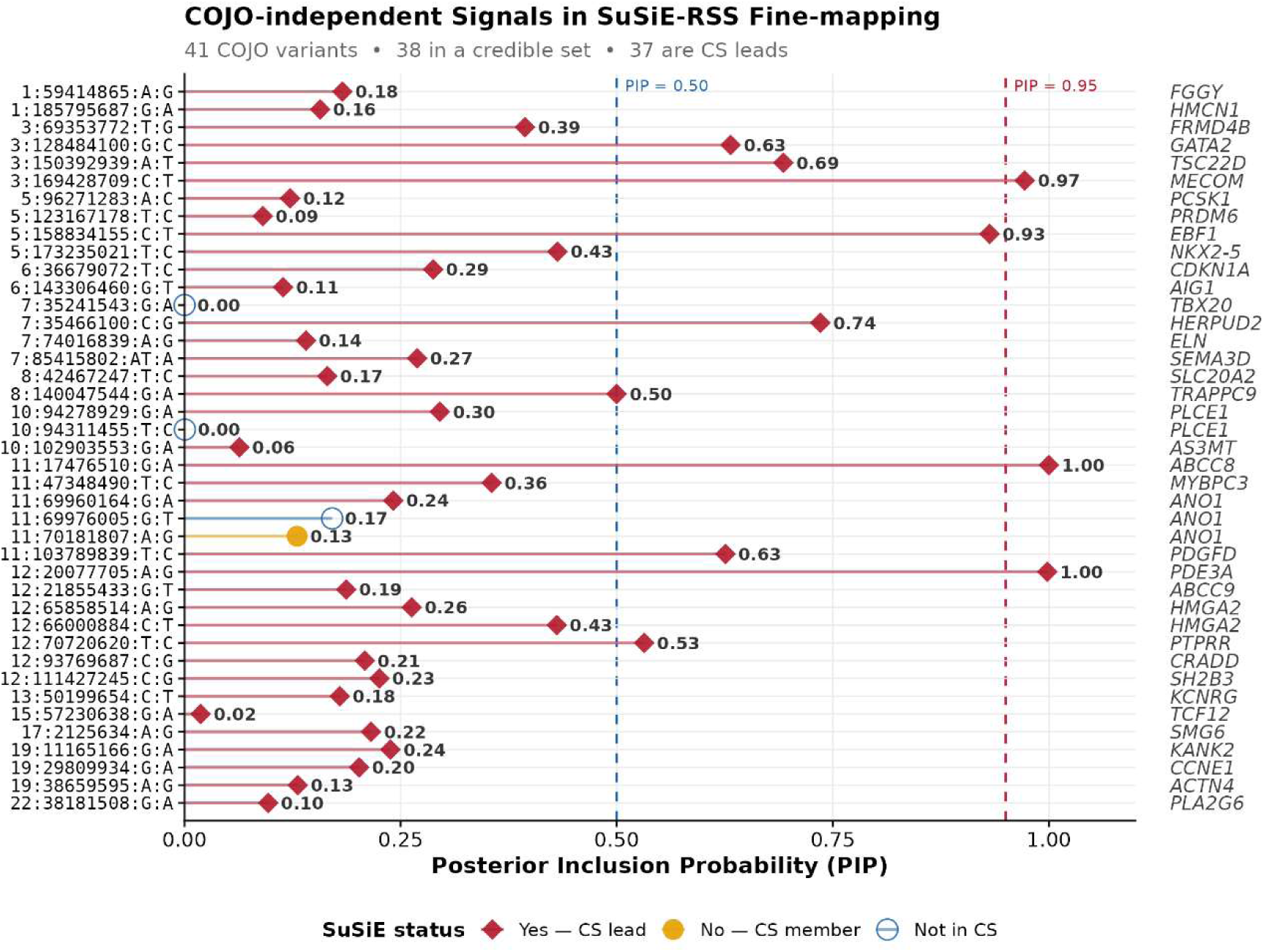
Statistical fine-mapping of PA diameter association signals using SuSiE-RSS. Lollipop plot of posterior inclusion probabilities (PIPs) from SuSiE-RSS fine-mapping of the 41 conditionally independent pulmonary artery (PA) diameter association signals identified by GCTA-COJO. Each row represents a COJO lead variant (left axis, chr:position:alleles), with the nearest protein-coding gene shown on the right axis. Point shape and color indicate credible set (CS) membership: red-filled diamonds indicate credible set lead variants, yellow-filled diamonds indicate non-lead credible set members, and open circles indicate variants not assigned to a credible set. PIP values are labeled next to each point. The blue dashed line marks PIP = 0.50, and the red dashed line marks PIP = 0.95.

Four missense variants were identified across three credible sets. A *MYBPC3* missense (Ser→Gly; PIP=0.36) was the credible-set lead at the chromosome 11 *MYBPC3*/*SLC39A13* locus, while *PLCE1* (Arg→Pro; PIP=0.14) and *SH2B3* (Trp→Arg; PIP=0.19) missense variants were within but not the lead of their respective credible sets. A complete annotation summary is provided in the Supplementary Results (Table S10).

### Colocalization of PA Diameter Association Signals with Lung and Cardiovascular Tissue eQTLs Prioritizes Candidate Effector Genes

Second, we performed Bayesian colocalization analyses with eQTLs in cardiopulmonary-relevant tissues, finding shared causal variation (PP.H4≥0.8) at 5 association signals, nominating *ABCC8* (aorta, coronary artery), *PDGFD* (aorta), *HMCN1* (tibial artery), *CCNE1* (heart atrial appendage), and *SMIM19* (coronary artery) as candidate effector genes (**Figure 5**).

**Figure 5.**
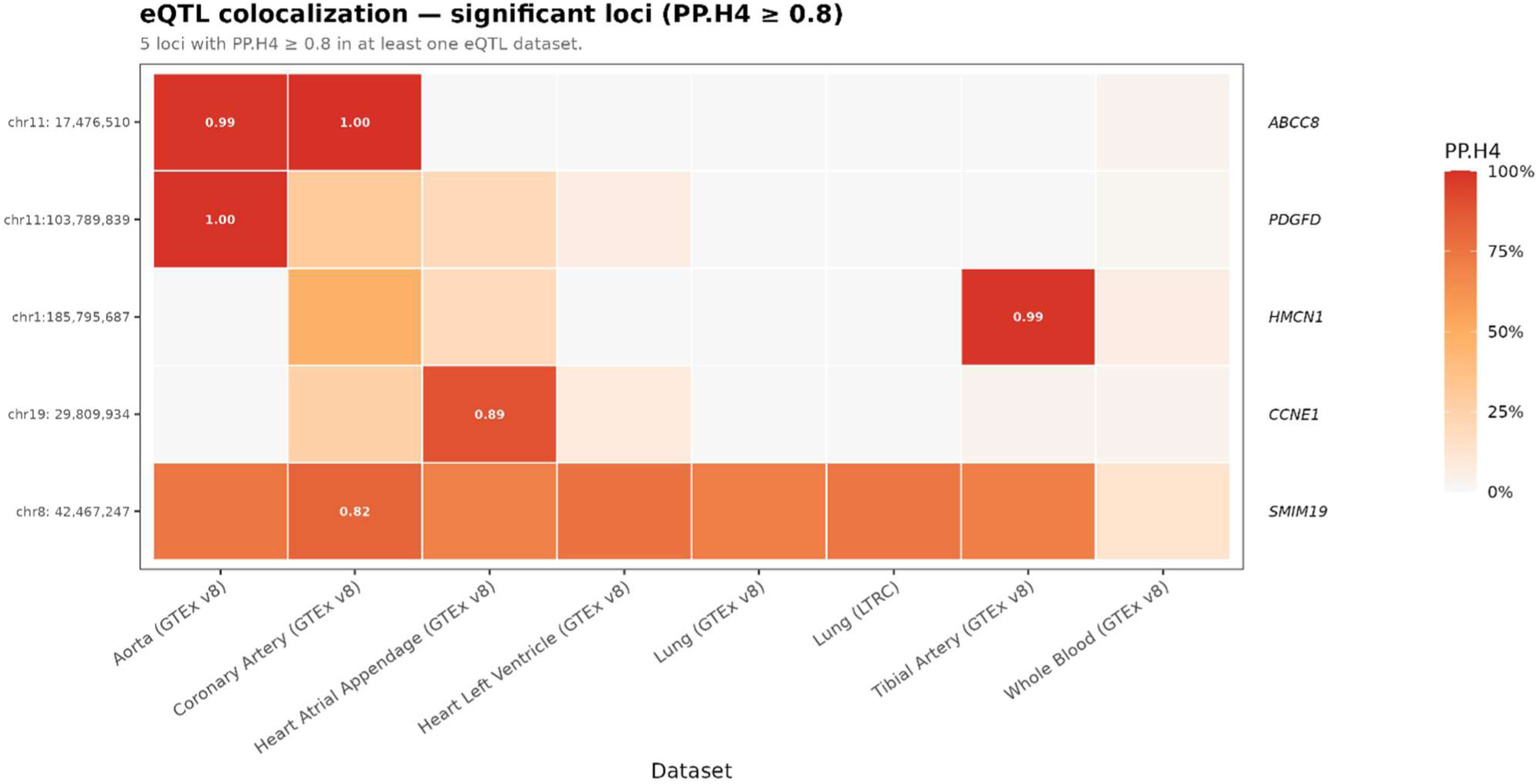
Colocalization of pulmonary artery diameter association signals with tissue-specific eQTLs. Heatmap of Bayesian colocalization posterior probabilities (PP.H4) between the 41 conditionally independent pulmonary artery (PA) diameter association signals and seven tissue-specific eQTL datasets. Rows represent independent PA diameter signals, and columns represent eQTL datasets from lung (Lung Tissue Research Consortium), coronary artery, tibial artery, heart atrial appendage, heart left ventricle and whole blood (GTEx v8). Cells are colored by the posterior probability of a shared causal variant (PP.H4) from coloc.abf analysis, with PP.H4 ≥ 0.8 indicating strong evidence of colocalization. Candidate effector genes prioritized through eQTL colocalization are annotated alongside the corresponding PA diameter signals.

### Colocalization of PA Diameter Signals with Pulse Pressure Reveals Shared Vascular Genetic Architecture

To further assess pleiotropic vascular effects, we performed colocalization with GWAS for pulse pressure and pulmonary arterial hypertension. Pulse pressure GWAS demonstrated the strongest overlap, with 15 PA diameter signals showing evidence of shared causal variation (Figure S7), indicating substantial shared genetic architecture between pulmonary vascular structure and systemic vascular biology. Several of these loci also colocalized with tissue-specific eQTLs, further supporting regulatory mechanisms linking PA structure to broader vascular phenotypes. Representative regional colocalization patterns between PA GWAS and signals at colocalized loci is shown in Figure S8.

### Integrated Genetic and Regulatory Evidence Prioritizes Candidate Effector Genes at PA Diameter Loci

To identify additional evidence for effector genes, we queried lead variants in Open Targets Genetics and harmonized from study-specific labels into nine biologically coherent domains (Table S11). For each association signal, candidate effector genes were prioritized by integrating multiple complementary lines of evidence (**Figure 6**; per-signal detail in Table S12). The prioritized gene matched the nearest protein-coding gene at 35 of 41 tested signals (85%). Prioritization was driven predominantly by regulatory and colocalization evidence, with qualifying coding variants contributing at three signals *(MYBPC3, PLCE1, SH2B3*). Twelve signals showed at least three converging layers (including *HMCN1, ABCC8,* and *CCNE1* with four layers), and six had no qualifying layer above thresholds. Six prioritized genes differed from the nearest positional candidate, underscoring the value of integrative gene prioritization beyond proximity-based annotation. Representative regional association plots are shown in **Figure 7**; the remaining loci are shown in Figure S9.

**Figure 6.**
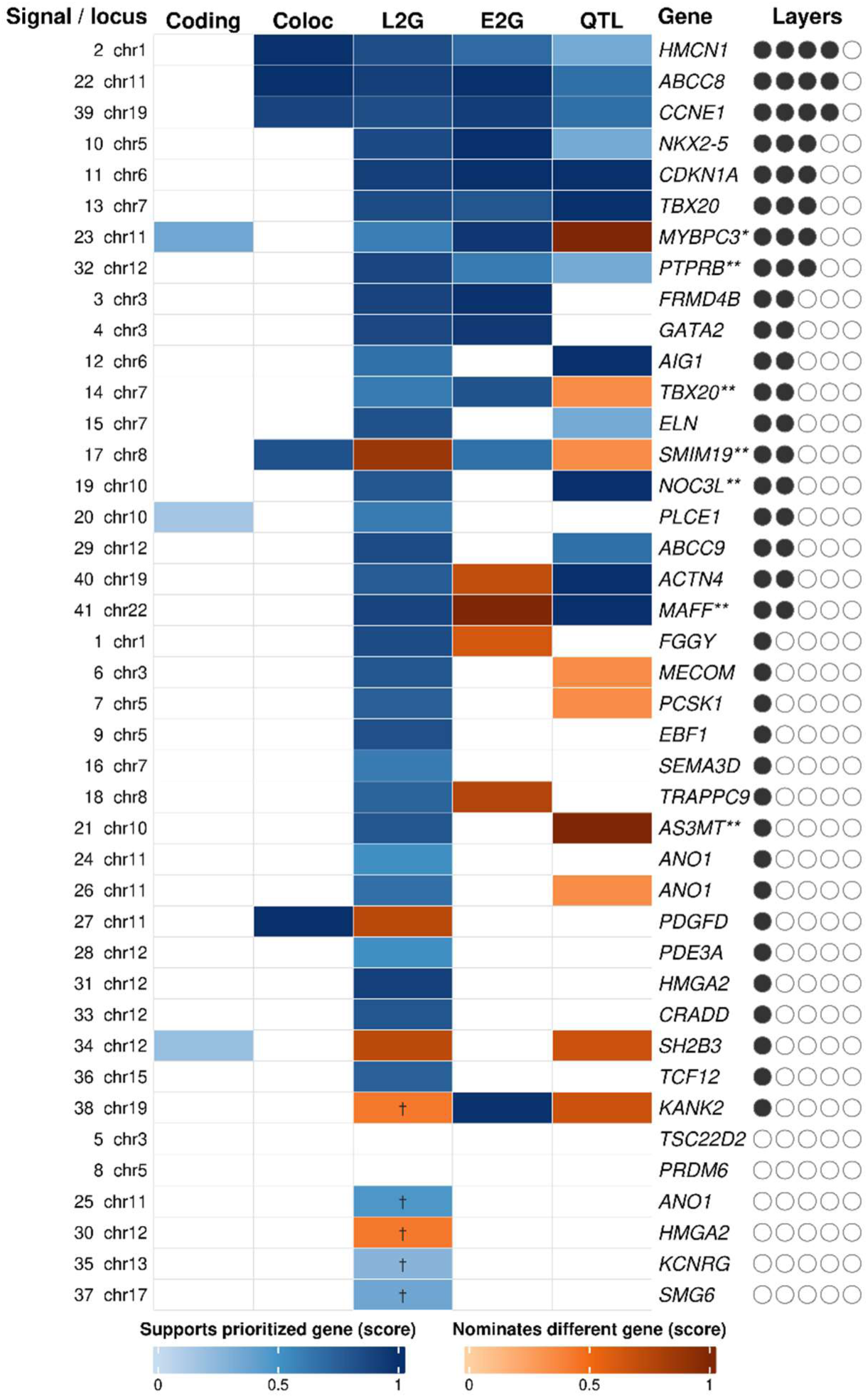
Integrated effector-gene prioritization across the 41 PA diameter association signals. Heatmap representation of the five-source effector-gene prioritization framework applied to the 41 independent pulmonary artery (PA) diameter association signals. Rows represent association signals, ordered by the number of convergent evidence (descending). Columns represent the five evidence sources, shown in prioritization order: Coding (credible-set coding variant, posterior inclusion probability (PIP) ≥ 0.10), Coloc (eQTL colocalization with GTEx v8 tissues, PP.H4 ≥ 0.80), L2G (Open Targets Genetics locus-to-gene score), E2G (Open Targets enhancer-to-gene linkage score), and QTL (Open Targets molecular QTL evidence, including eQTL, pQTL, sQTL, scEQTL, and tuQTL). Cell color: intensity reflects the strength of evidence on a 0–1 scale (PIP for Coding; PP.H4 for Coloc; score for L2G and E2G, and number of QTL modalities normalized to 3 for QTL). Blue cells indicate agreement with the prioritized effector gene at that signal. Orange cells indicate nomination of a different gene, highlighting discordance among evidence sources. White cells indicate missing evidence layer. A dagger (†) over a cell marks an L2G or E2G nomination shown for context but below its high-confidence threshold; such cells are not counted toward the converging-layers circles. Right-side annotation: the prioritized effector gene per signal, with a double asterisk (**) when the prioritized gene differs from the nearest protein-coding gene (eight signals), and a single asterisk (*) marking the row in which the coding variant is itself the credible-set lead variant (*MYBPC3*). Filled circles show the number of evidence layers above the high-confidence thresholds, that is, the converging-layers count (maximum five). Thresholds: L2G ≥ 0.50, E2G ≥ 0.60, Coding PIP ≥ 0.10, Coloc PP.H4 ≥ 0.80, and QTL evidence from any modality. Left-side annotation: COJO signal number and chromosome.

**Figure 7.**
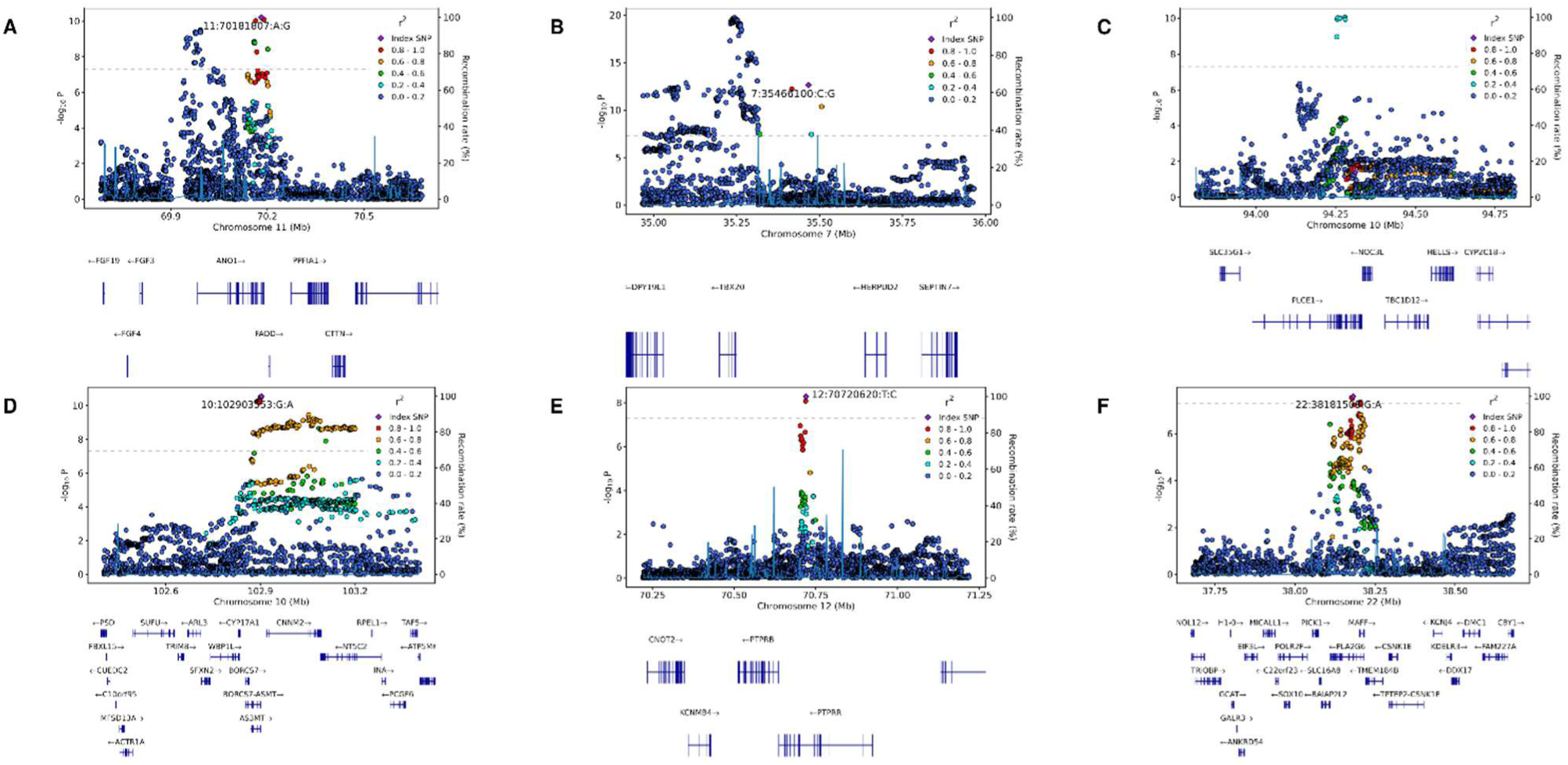
Selected regional association (LocusZoom) plots illustrating multiple signals near *ANO1* and the reassignment of effector genes by integrative annotation. Each panel displays regional association plots showing −log₁₀ association P values for PA diameter (left axis) across genomic position, with recombination rate overlaid (right axis). The lead variant is shown as a purple diamond, and surrounding variants are colored by linkage disequilibrium (r²) with the lead variant. Gene models are shown below each panel. (A) The *ANO1* locus on chromosome 11, shown for one of its three conditionally independent signals. A second, LD-distinct association peak is visible within the same region, illustrating allelic heterogeneity at the locus and supporting multiple independent associations converging on *ANO1*. (B–E) Loci at which integrative gene prioritization identified an effector gene different from the nearest protein-coding gene, highlighting the limitations of proximity-based annotation in gene-dense regions: (B) *TBX20* over the nearer *HERPUD2* (chromosome 7); (C) *NOC3L* over *PLCE1* (chromosome 10); (D) *AS3MT* over *BORCS7-ASMT* (chromosome 10); (E) *PTPRB* over *PTPRR* (chromosome 12) and (F) *MAFF* over *PLA2G6* (chromosome 22)

Open Targets–native summaries of L2G score against maximum fine-mapping posterior probability (Figure S10) and of multi-modality QTL convergence (Figure S11) independently placed *ABCC8* among the highest-confidence genes, corroborating the integrated prioritization. pQTL association results from the UK Biobank Pharma Proteomics Project and cell specific expression patterns in the Human Lung CellRef single-cell atlas are presented in Table S13 and Figure S12.

## DISCUSSION

In the largest genetic study of pulmonary artery diameter to date, encompassing more than 51,000 participants from COPD-enriched and population-based cohorts, we identified 44 independent association signals across 39 loci, expanding the genetic landscape of pulmonary vascular structure. Despite marked differences in smoking exposure, COPD burden, and imaging modality, genetic effects were highly concordant across cohorts, supporting a shared biological basis of pulmonary artery size. Integrative fine-mapping and regulatory annotation prioritized candidate effector genes involved in vascular remodeling, ion-channel signaling, and cardiopulmonary development, providing mechanistic insights beyond locus discovery.

*ANO1* emerged as one of the most compelling signals, supported by genome-wide significance in the COPD-enriched discovery cohort and three independent association signals in the larger meta-analysis. *ANO1* (TMEM16A) encodes a calcium-activated chloride channel expressed in pulmonary arterial smooth muscle cells, where it contributes to membrane depolarization, vasoconstriction, and vascular remodeling (41). The presence of multiple independent signals suggests that distinct regulatory elements may converge on smooth muscle excitability and remodeling. However, credible-set variants extending into an unannotated genomic region at the locus complicate effector-gene prioritization, despite the strong biological candidacy.

Bayesian fine-mapping identified several high-confidence variants near *MECOM*, *ABCC8*, and *PDE3A*, nominating these variants as plausible candidates for functional validation. The variable resolution across loci, reflecting differences in local LD, effect size, and ancestry, underscores the value of integrating fine-mapping with orthogonal regulatory evidence. Functional annotation of the credible-set variants reinforced a predominantly regulatory architecture. Even where coding variants were present, they rarely drove the signal: only a *MYBPC3* Ser→Gly variant emerged as credible-set lead (PIP=0.36), whereas missense variants in *PLCE1* (Arg→Pro) and *SH2B3* (Trp→Arg) showed lower posterior probability. Notably, the most confidently fine-mapped variant in the study was a promoter variant in *ABCC8* resolved to a single-variant credible set (PIP=1.00). Additional high-confidence regulatory candidates localized to the promoter of *FRMD4B* and the 3’ untranslated region of *KANK2*, while several highly probable promoter variants mapped to uncharacterized non-coding transcripts. Together with the credible-set variants extending into an unannotated region at *ANO1*, these findings highlight the importance of integrating fine-mapping with regulatory annotation to resolve effector genes and provide variant-level evidence that common regulatory, rather than protein-altering, variation predominantly shapes pulmonary artery structure.

Colocalization with tissue eQTLs nominated candidate effector genes in cardiovascular tissues, including *ABCC8, PDGFD, HMCN1*, and *CCNE1,* linking common regulatory variation at these loci to gene expression in arteries and the heart. Colocalization with pulse pressure identified 15 shared signals, demonstrating substantial overlap between the genetic determinants of pulmonary vascular structure and systemic vascular biology and implicating shared pathways governing vascular tone, remodeling, and arterial stiffness whereas overlap with PAH was limited.

Integrating additional layers of evidence refined candidate genes and exposed the limits of proximity-based assignment. At six signals the prioritized effector differed from the nearest protein-coding gene, most notably prioritizing *TBX20* over the adjacent *HERPUD2* and *SMIM19* over *SLC20A2*, with prioritized genes converging on vascular tone regulation, smooth muscle proliferation, extracellular matrix remodeling, and cardiopulmonary development (13,42–45). Ion channel (*ABCC8, KCNRG and ANO1*) and second messenger signaling (*PDE3A*) featured prominently*. PDGFD* further implicates growth-factor–responsive remodeling and has itself been proposed as an idiopathic PAH risk gene (44,46,47), while *TBX20*, *HMCN1*, *and CCNE1* map to developmental, structural, and proliferative pathways (48–50). *ABCC8* provides the clearest example of convergence between rare and common genetic architecture. Rare loss-of-function variants in *ABCC8* cause heritable PAH (45), whereas the common regulatory variant identified here is associated with PA diameter in the general population. This observation suggests that rare disease-causing and common susceptibility variants can act through shared biological pathways governing pulmonary vascular structure.

Several limitations warrant consideration. The independent replication sample was relatively modest, and larger studies are needed to validate additional loci. Rare-variant analyses suggested that large-effect rare coding variation is not the dominant contributor to variation in PA traits, as only *GEM* reached exome-wide significance; however, as larger sequencing cohorts with deep cardiopulmonary phenotyping become available, future studies may further clarify the interplay between rare and common genetic determinants of pulmonary vascular traits. The limited number of eQTL colocalization likely reflects the constraints of currently available bulk-tissue; variants influencing PA size may act within specialized pulmonary vascular cell populations (smooth muscle cells, pericytes, and specialized endothelial subtypes) that are underrepresented in bulk lung and vascular tissue datasets. Consistent with this, prioritized genes were enriched in these populations in the Human Lung CellRef atlas, highlighting the need for cell-type–specific regulatory maps of the pulmonary vasculature to resolve the mechanisms linking genetic variation to remodeling. The study was not powered to detect disease- or exposure-specific genetic effects, including gene-smoking interactions. Variant-to-gene mapping through Open Targets relied on overlaps with existing credible sets rather than comprehensive re-analysis of primary datasets. Despite these limitations, convergent prioritization across multiple orthogonal datasets provides a strong foundation for future mechanistic and translational studies.

## CONCLUSION

By integrating whole-genome sequencing in COPD-enriched cohorts with imputed-genotype analyses in population-based imaging cohorts, we identified multiple loci associated with PA size and prioritized candidate effector genes through coding-variant annotation, colocalization, and regulatory evidence. Because PA diameter is readily measurable on routine chest imaging and exhibits remarkable genetic consistency across diverse populations and imaging approaches, our results establish PA structure as a heritable and genetically informative imaging endophenotype for pulmonary vascular disease and provide a framework for mechanistic investigation, risk stratification, and translational study in pulmonary hypertension.

## Data availability

Whole genome sequencing and phenotype data for COPDGene is available through dbGaP Study Accession: phs000179.v7.p2; for ECLIPSE phs001252.v1.p1; and for Framingham: phs000007.v34.p15; summary statistics are available through TOPMed (phs001974).

## Author’s contribution

Conception and design: V.F., E.K.S, A.B., M.H.C. Analysis and interpretation: V.F., K.K., C.T., Y.Q., Y.J., G.W., R.C.W., J.M.W., H.L., G.T.O., E.K.S., A.V.S, S.B.G, N.G, E.K.S., A.B., and M.H.C. All authors were involved in drafting the manuscript and contributed important intellectual content.

## ACKNOWLEDGMENTS

Molecular data for the Trans-Omics in Precision Medicine (TOPMed) program was supported by the National Heart, Lung and Blood Institute (NHLBI). Whole-genome sequencing for NHLBI TOPMed studies was performed at participating TOPMed sequencing centers. Core support, including centralized genomic read mapping, genotype calling, variant quality metrics, and filtering were provided by the TOPMed Informatics Research Center (3R01HL-117626-02S1; contract HHSN268201800002I). Additional core support, including phenotype harmonization, data management, sample-identity QC, and program coordination, was provided by the TOPMed Data Coordinating Center (R01HL-120393; U01HL-120393; contract HHSN268201800001I). We gratefully acknowledge the studies and participants who provided biological samples and data for the TOPMed program.

The COPDGene project described was supported by Award Number U01 HL089897 and Award Number U01 HL089856 from the National Heart, Lung, and Blood Institute. The content is solely the responsibility of the authors and does not necessarily represent the official views of the National Heart, Lung, and Blood Institute or the National Institutes of Health. The COPDGene project is also supported by the COPD Foundation through contributions made to an Industry Advisory Board comprised of AstraZeneca, Boehringer Ingelheim, GlaxoSmithKline, Novartis, Pfizer, Siemens and Sunovion.

## COPDGene^®^ Investigators – Core Units

*Administrative Center*: James D. Crapo, MD (PI); Edwin K. Silverman, MD, PhD (PI); Barry J. Make, MD; Elizabeth A. Regan, MD, PhD

*Genetic Analysis Center*: Terri H. Beaty, PhD; Peter J. Castaldi, MD, MSc; Michael H. Cho, MD, MPH; Dawn L. DeMeo, MD, MPH; Adel El Boueiz, MD, MMSc; Marilyn G. Foreman, MD, MS; Auyon Ghosh, MD; Lystra P. Hayden, MD, MMSc; Craig P. Hersh, MD, MPH; Jacqueline Hetmanski, MS; Brian D. Hobbs, MD, MMSc; John E. Hokanson, MPH, PhD; Wonji Kim, PhD; Nan Laird, PhD; Christoph Lange, PhD; Sharon M. Lutz, PhD; Merry-Lynn McDonald, PhD; Dmitry Prokopenko, PhD; Matthew Moll, MD, MPH; Jarrett Morrow, PhD; Dandi Qiao, PhD; Elizabeth A. Regan, MD, PhD; Aabida Saferali, PhD; Phuwanat Sakornsakolpat, MD; Edwin K. Silverman, MD, PhD; Emily S. Wan, MD; Jeong Yun, MD, MPH

*Imaging Center*: Juan Pablo Centeno; Jean-Paul Charbonnier, PhD; Harvey O. Coxson, PhD; Craig J. Galban, PhD; MeiLan K. Han, MD, MS; Eric A. Hoffman, Stephen Humphries, PhD; Francine L. Jacobson, MD, MPH; Philip F. Judy, PhD; Ella A. Kazerooni, MD; Alex Kluiber; David A. Lynch, MB; Pietro Nardelli, PhD; John D. Newell, Jr., MD; Aleena Notary; Andrea Oh, MD; Elizabeth A. Regan, MD, PhD; James C. Ross, PhD; Raul San Jose Estepar, PhD; Joyce Schroeder, MD; Jered Sieren; Berend C. Stoel, PhD; Juerg Tschirren, PhD; Edwin Van Beek, MD, PhD; Bram van Ginneken, PhD; Eva van Rikxoort, PhD; Gonzalo Vegas Sanchez-Ferrero, PhD; Lucas Veitel; George R. Washko, MD; Carla G. Wilson, MS;

*PFT QA Center, Salt Lake City, UT*: Robert Jensen, PhD

*Data Coordinating Center and Biostatistics*, *National Jewish Health, Denver, CO*: Matthew Strand, PhD; Jim Crooks, PhD; Katherine Pratte, PhD; Aastha Khatiwada, PhD; Carla G. Wilson, MS

*Epidemiology Core*, *University of Colorado Anschutz Medical Campus, Aurora, CO*: John E. Hokanson, MPH, PhD; Erin Austin, PhD; Gregory Kinney, MPH, PhD; Sharon M. Lutz, PhD; Kendra A. Young, PhD

*Mortality Adjudication Core:* Surya P. Bhatt, MD; Jessica Bon, MD; Alejandro A. Diaz, MD, MPH; MeiLan K. Han, MD, MS; Barry Make, MD; Susan Murray, ScD; Elizabeth Regan, MD; Xavier Soler, MD; Carla G. Wilson, MS

*Biomarker Core*: Russell P. Bowler, MD, PhD; Katerina Kechris, PhD; Farnoush Banaei-Kashani, PhD

## COPDGene^®^ Investigators – Clinical Centers

*Ann Arbor VA:* Jeffrey L. Curtis, MD; Perry G. Pernicano, MD

*Baylor College of Medicine, Houston, TX*: Nicola Hanania, MD, MS; Mustafa Atik, MD; Aladin Boriek, PhD; Kalpatha Guntupalli, MD; Elizabeth Guy, MD; Amit Parulekar, MD;

*Brigham and Women’s Hospital, Boston, MA*: Dawn L. DeMeo, MD, MPH; Craig Hersh, MD, MPH; Francine L. Jacobson, MD, MPH; George Washko, MD

*Columbia University, New York, NY*: R. Graham Barr, MD, DrPH; John Austin, MD; Belinda D’Souza, MD; Byron Thomashow, MD

*Duke University Medical Center, Durham, NC*: Neil MacIntyre, Jr., MD; H. Page McAdams, MD; Lacey Washington, MD

*HealthPartners Research Institute, Minneapolis, MN*: Charlene McEvoy, MD, MPH; Joseph Tashjian, MD

*Johns Hopkins University, Baltimore, MD*: Robert Wise, MD; Robert Brown, MD; Nadia N. Hansel, MD, MPH; Karen Horton, MD; Allison Lambert, MD, MHS; Nirupama Putcha, MD, MHS

*Lundquist Institute for Biomedical Innovation at Harbor UCLA Medical Center, Torrance, CA*: Richard Casaburi, PhD, MD; Alessandra Adami, PhD; Matthew Budoff, MD; Hans Fischer, MD; Janos Porszasz, MD, PhD; Harry Rossiter, PhD; William Stringer, MD

*Michael E. DeBakey VAMC, Houston*, *TX*: Amir Sharafkhaneh, MD, PhD; Charlie Lan, DO

*Minneapolis VA:* Christine Wendt, MD; Brian Bell, MD; Ken M. Kunisaki, MD, MS

*Morehouse School of Medicine, Atlanta, GA*: Eric L. Flenaugh, MD; Hirut Gebrekristos, PhD; Mario Ponce, MD; Silanath Terpenning, MD; Gloria Westney, MD, MS

*National Jewish Health, Denver, CO*: Russell Bowler, MD, PhD; David A. Lynch, MB

*Reliant Medical Group, Worcester, MA*: Richard Rosiello, MD; David Pace, MD

*Temple University, Philadelphia, PA:* Gerard Criner, MD; David Ciccolella, MD; Francis Cordova, MD; Chandra Dass, MD; Gilbert D’Alonzo, DO; Parag Desai, MD; Michael Jacobs, PharmD; Steven Kelsen, MD, PhD; Victor Kim, MD; A. James Mamary, MD; Nathaniel Marchetti, DO; Aditi Satti, MD; Kartik Shenoy, MD; Robert M. Steiner, MD; Alex Swift, MD; Irene Swift, MD; Maria Elena Vega-Sanchez, MD

*University of Alabama, Birmingham, AL:* Mark Dransfield, MD; William Bailey, MD; Surya P. Bhatt, MD; Anand Iyer, MD; Hrudaya Nath, MD; J. Michael Wells, MD

*University of California, San Diego, CA*: Douglas Conrad, MD; Xavier Soler, MD, PhD; Andrew Yen, MD

*University of Iowa, Iowa City, IA*: Alejandro P. Comellas, MD; Karin F. Hoth, PhD; John Newell, Jr., MD; Brad Thompson, MD

*University of Michigan, Ann Arbor, MI:* MeiLan K. Han, MD MS; Ella Kazerooni, MD MS; Wassim Labaki, MD MS; Craig Galban, PhD; Dharshan Vummidi, MD

*University of Minnesota, Minneapolis, MN*: Joanne Billings, MD; Abbie Begnaud, MD; Tadashi Allen, MD

*University of Pittsburgh, Pittsburgh, PA*: Frank Sciurba, MD; Jessica Bon, MD; Divay Chandra, MD, MSc; Joel Weissfeld, MD, MPH

*University of Texas Health, San Antonio, San Antonio, TX*: Antonio Anzueto, MD; Sandra Adams, MD; Diego Maselli-Caceres, MD; Mario E. Ruiz, MD; Harjinder Singh

The ECLIPSE study (NCT00292552) was sponsored by GlaxoSmithKline. The ECLIPSE investigators included: ECLIPSE Investigators — Bulgaria: Y. Ivanov, Pleven; K. Kostov, Sofia. Canada: J. Bourbeau, Montreal; M. Fitzgerald, Vancouver, BC; P. Hernandez, Halifax, NS; K. Killian, Hamilton, ON; R. Levy, Vancouver, BC; F. Maltais, Montreal; D. O’Donnell, Kingston, ON. Czech Republic: J. Krepelka, Prague. Denmark: J. Vestbo, Hvidovre. The Netherlands: E. Wouters, Horn-Maastricht. New Zealand: D. Quinn, Wellington. Norway: P. Bakke, Bergen. Slovenia: M. Kosnik, Golnik. Spain: A. Agusti, J. Sauleda, P. de Mallorca. Ukraine: Y. Feschenko, V. Gavrisyuk, L. Yashina, Kiev; N. Monogarova, Donetsk. United Kingdom: P. Calverley, Liverpool; D. Lomas, Cambridge; W. MacNee, Edinburgh; D. Singh, Manchester; J. Wedzicha, London. United States: A. Anzueto, San Antonio, TX; S. Braman, Providence, RI; R. Casaburi, Torrance CA; B. Celli, Boston; G. Giessel, Richmond, VA; M. Gotfried, Phoenix, AZ; G. Greenwald, Rancho Mirage, CA; N. Hanania, Houston; D. Mahler, Lebanon, NH; B. Make, Denver; S. Rennard, Omaha, NE; C. Rochester, New Haven, CT; P. Scanlon, Rochester, MN; D. Schuller, Omaha, NE; F. Sciurba, Pittsburgh; A. Sharafkhaneh, Houston; T. Siler, St. Charles, MO; E. Silverman, Boston; A. Wanner, Miami; R. Wise, Baltimore; R. ZuWallack, Hartford, CT. ECLIPSE Steering Committee: H. Coxson (Canada), C. Crim (GlaxoSmithKline, USA), L. Edwards (GlaxoSmithKline, USA), D. Lomas (UK), W. MacNee (UK), E. Silverman (USA), R. Tal-Singer (Co-chair, GlaxoSmithKline, USA), J. Vestbo (Co-chair, Denmark), J. Yates (GlaxoSmithKline, USA). ECLIPSE Scientific Committee: A. Agusti (Spain), P. Calverley (UK), B. Celli (USA), C. Crim (GlaxoSmithKline, USA), B. Miller (GlaxoSmithKline, USA), W. MacNee (Chair, UK), S. Rennard (USA), R. Tal-Singer (GlaxoSmithKline, USA), E. Wouters (The Netherlands), J. Yates (GlaxoSmithKline, USA).

