## Supplementary material for "Genetic Determinants of Pulmonary Artery Size in over 50,000 Subjects with and without COPD": Online Supplement (Figures S1-S12)

### Contents

#### Supplementary Methods

1. Cohorts and phenotype definitions
2. Common- and rare-variant association framework
3. Meta-analysis
4. Secondary phenotype (PA/A ratio)
5. Replication in the Framingham Heart Study
6. Definition of genomic risk loci

7. Conditional and joint analysis
8. Statistical fine-mapping
9. Predicted molecular consequence of credible-set variants
10. Heritability and genetic correlation
11. Colocalization
12. Open Targets Genetics annotation and phenotype harmonization
13. Evidence integration and effector-gene prioritization
14. Single-cell expression

**Supplementary Results**

1. Rare-variant gene-based analysis
2. Secondary phenotype (PA/A ratio)
3. Heritability and genetic correlation
4. Fine-mapping and credible-set annotation
5. Colocalization
6. Open Targets prioritization
7. Cellular expression of prioritized effector genes

**Supplementary Figures (Figures S1–S12)**

**Supplementary Table Legends (Tables S1–S13)**

**Supplementary References**

---

### Supplementary Methods

#### 1. Cohorts and phenotype definitions

The **Genetic Epidemiology of COPD (COPDGene)** study is a multi-institutional NHLBI-funded cohort designed to investigate the epidemiologic and genomic determinants of COPD (1). The study enrolled 10,198 current and former smokers aged 45 to 80 years with at least 10 pack-years of lifetime smoking who self-identified as non-Hispanic White or African American. At each five-year study visit, participants underwent comprehensive longitudinal phenotyping, including spirometry, questionnaires, and inspiratory/expiratory chest CT scans, performed using standardized protocols with centralized quality control. 5-year follow-up (Phase 2) was completed in 6,153 subjects. The maximal main pulmonary artery (PA) diameter was measured at the level of the PA bifurcation in both Phase 1 and Phase 2 by trained readers blinded to clinical characteristics (2).

The **Evaluation of COPD Longitudinally to Identify Predictive Surrogate Endpoints (ECLIPSE)** study is a longitudinal observational cohort that enrolled 2,164 White subjects with COPD, 337 smoking controls, and 245 nonsmoking controls and followed them for three years (3). Inclusion criteria included age 40-75 years, a smoking history of at least 10 pack-years of smoking, and spirometry consistent with GOLD grades 2-4 (cases) or normal spirometry (controls). Study visits were conducted at enrollment, three months, and every six months thereafter, with serial spirometry, questionnaires, and standardized clinical assessments performed at each visit. Chest CT scans were acquired at baseline, one year, and three years. Maximal main PA diameter was measured on baseline CT scans by the same trained readers who performed PA measurements in COPDGene. Whole-genome sequencing data (Freeze 10) for COPDGene and ECLIPSE were obtained through the NHLBI Trans-Omics in Precision Medicine (TOPMed) program (4).

The **UK Biobank** is a large population-based cohort of more than 500,000 participants recruited at ages 40-69 years (5). Cardiac MRI was performed using standardized imaging protocols across participating centers. The main PA was segmented from cine MRI images using a deep learning model trained on manually annotated scans, generating multiple quantitative traits including PA root diameter and systolic and diastolic PA dimensions (6). We analyzed systolic PA diameter, as it most closely corresponds to CT-based PA measurements (7). We obtained publicly available GWAS summary statistics from the Cardiovascular Disease Knowledge Portal (CVDKP, https://cvd.hugeamp.org/).

The **Framingham Heart Study (FHS)** is a community-based longitudinal cohort to investigate the genetic and environmental determinants of cardiovascular disease through repeated standardized examinations. FHS served as an independent replication cohort, including participants from the Offspring and Third Generation cohorts evaluated between 2002 and 2005. Participants underwent non-contrast chest CT, enabling high-resolution characterization of thoracic vascular anatomy. The maximum main PA diameter was measured on axial CT images at the level of the right PA using standardized protocols (8). GWAS summary statistics were provided by investigators at the University of Massachusetts Chan Medical School.

**2. Common- and rare-variant association framework**

For the combined COPDGene and ECLIPSE cohort, GWAS were performed using 11,927,683 high-quality common variants (MAF ≥ 0.01) derived from whole-genome sequencing following standard variant-level quality control and mapped to GRCh38.

Association analyses were conducted using REGENIE v4 (9), a whole-genome regression framework that accounts for population structure and relatedness. Ridge regression models based on LD-pruned common variants selected with PLINK 2 (10) were used to generate polygenic prediction terms, followed by association testing for common and rare variants using the same covariates: age, sex, height, study cohort, sequencing center, smoking pack-years, $FEV_1$, body mass index, and the first 20 genetic ancestry principal components, with mean-centered squared terms for age and height added to capture nonlinear effects. Secondary analyses included PA/A as a continuous trait and PA enlargement (PA/A > 1). Firth logistic regression was applied for the binary PA enlargement (PA/A > 1) trait to mitigate sparse-data bias (11). Additional exploratory analyses were performed to assess robustness and heterogeneity of genetic effects, including cohort-specific analyses in COPDGene and ECLIPSE separately, independent evaluation in COPDGene Phase 2, and stratified analyses by COPD status.

Rare variants (MAF < 0.01) were assessed using single-variant and gene-based burden tests across two functional masks defined by predicted protein impact (12). Genome-wide and exome-wide significance were defined as $P < 5 \times 10^{-8}$ and $P < 2.5 \times 10^{-6}$, respectively, the latter reflecting correction for the multiple-testing burden of gene-based rare-variant analyses. Gene-based analyses were performed across aggregate allele frequency thresholds of ≤0.1, ≤0.01, and ≤0.001, with additional sensitivity analyses requiring a minimum number of minor alleles. Two variant masks were used: (1) putative loss-of-function (pLOF) variants, defined as stop_gained, start_lost, splice_donor_variant, splice_acceptor_variant, stop_lost, or frameshift_variant annotations; and (2) a broader deleterious mask combining pLOF variants with high-confidence damaging missense variants. Missense variants were classified as damaging only if all five algorithms (SIFT, PolyPhen-2 HDIV, PolyPhen-2 HVAR, LRT, and MutationTaster) predicted a deleterious effect (13). Functional annotation was performed using Whole Genome Sequence Annotator (WGSA) v0.95 (14) and the Ensembl Variant Effect Predictor (VEP) (15).

**3. Meta-analysis**

Stage 2 and Stage 3 meta-analyses were performed using METAL (release 2011-03-25) (16). UK Biobank GWAS summary statistics were converted to GRCh38 coordinates using the UCSC LiftOver tool (17) (Lift Genome Annotations). The Stage 2 secondary analysis of PA/A incorporated all cohorts with available aortic measurements in COPDGene, ECLIPSE, and the UK Biobank.

**4. Additional Pulmonary Artery Phenotypes: PA/A ratio and PA Enlargement**

In addition to absolute PA diameter, we analysed the pulmonary-artery-to-ascending-aorta (PA/A) ratio as a secondary continuous phenotype and PA enlargement (PA/A > 1) as a binary phenotype. The ascending aorta was measured at the same axial level as the main PA in each cohort. Both phenotypes were tested using the same

REGENIE framework, covariates, and quality-control procedures applied to PA diameter, with Firth logistic regression used for the binary PA-enlargement trait to mitigate sparse-data bias. Stage 1 discovery (COPDGene + ECLIPSE) and Stage 2 meta-analysis were performed analogously to the primary analysis; because the ratio requires an aortic measurement, the PA/A Stage 2 meta-analysis combined COPDGene, ECLIPSE, and the UK Biobank. Genome-wide significant PA/A loci were defined using the same ±500 kb window locus-definition strategy applied to PA diameter and are reported in Table S5. Cross-cohort effect-size concordance for PA/A was evaluated using the same approach as for PA diameter (Figure S5), and SNP heritability and cross-cohort genetic correlation were estimated by LD score regression for both PA/A and PA diameter.

**5. Replication in the Framingham Heart Study**

The Framingham Heart Study contributed in two ways. First, FHS served as an independent replication cohort for the Stage 2 PA diameter lead variants: variants were harmonized to the discovery effect allele, with effect-allele frequencies checked for correct orientation, and replication was defined as a nominally significant association ($P < 0.05$) with a concordant direction of effect. Of the 37 Stage 2 lead variants, 36 were available in FHS, eight of which replicated at nominal significance, including four loci not previously associated with PA diameter (Table S7). Second, FHS was incorporated into the Stage 3 four-cohort joint meta-analysis, increasing the total sample size to 51,639 participants and contributing to the final set of loci carried forward to conditional analysis and fine-mapping.

### 6. Definition of genomic risk loci

Genome-wide significant variants ($P < 5\times10^{-8}$) were grouped into genomic risk loci using a ±500 kb window-merging strategy, with the most significant variant designated as the lead variant for each locus. For fine-mapping analyses, loci were similarly defined by applying the same ±500 kb window around each conditionally independent signal identified by GCTA-COJO.

### 7. Conditional and joint analysis

Conditionally independent PA association signals were identified using conditional and joint analysis (GCTA-COJO; LD threshold $r^2 < 0.9$) applied to Stage 3 results restricted to participants of self-reported European ancestry (18,19). Independent signals were selected using the stepwise model-selection procedure (cojo-slct) and subsequently confirmed by conditional analysis (cojo-cond).

### 8. Statistical fine-mapping

Fine-mapping loci were defined using a ±500 kb window-merging strategy centered on genome-wide significant variants. Stage 3 GWAS summary statistics were extracted for all variants within each locus and intersected with variants present in the self-reported European-ancestry COPDGene/ECLIPSE in-sample LD reference panel. Loci with fewer than 50 overlapping variants were excluded.

Per-locus signed Pearson correlation matrices were computed using PLINK2, and effect alleles in GWAS summary statistics were aligned to the LD reference panel. LD matrices were evaluated for positive semi-definiteness and regularized when necessary. Consistency between LD structure and GWAS summary statistics was assessed using estimate_s_rss(), with per-variant diagnostics generated using kriging_rss() to identify potential outliers.

Fine-mapping was performed using SuSiE-RSS (20,20) with a maximum of 10 causal signals per locus, 95% credible set coverage, and a minimum absolute correlation (purity) threshold of $|r| \geq 0.5$. Because the in-sample COPDGene/ECLIPSE reference panel was used, both residual and prior variances were estimated directly from the data. For each locus, 95% credible sets and posterior inclusion probabilities (PIPs) were computed, with lead variants defined as those with the highest PIP within each credible set.

**9. Predicted molecular consequence of credible-set variants**

Variants in 95% SuSiE-RSS credible sets were annotated for predicted molecular consequence in R using the VariantAnnotation package (21) against TxDb.Hsapiens.UCSC.hg38.knownGene on GRCh38. Coding consequences (missense, nonsense, synonymous) were assigned using predictCoding() with BSgenome.Hsapiens.UCSC.hg38, whereas non-coding feature overlap, including canonical splice site, 5′UTR, 3′UTR, promoter (2 kb upstream of the transcription start site), intron, and intergenic, were assigned using locateVariants() and AllVariants(). To generate a single annotation per variant, consequences were prioritized according to the following hierarchy: nonsense > missense > splice site > 5′UTR > 3′UTR > synonymous > promoter > intron > intergenic.

### 10. Heritability and genetic correlation

SNP-based heritability ($h^2$) of PA diameter and the PA/A ratio was estimated using LD score regression (LDSC) (22) via the LDscore module of the LDlink platform (23) (LDlink | Home), which implements the LD score regression method. Analyses used pre-computed LD scores derived from the 1000 Genomes Phase 3 European reference panel (24) and were restricted to HapMap3 variants (25). Heritability was estimated separately in the COPD-enriched (COPDGene + ECLIPSE) and UK Biobank cohorts to compare genetic architecture across the disease-enriched and population-based settings. Genetic correlations (rg) between the two settings were estimated using cross-trait LDSC (26). LDSC outputs included the total observed-scale $h^2$ with standard error, $\lambda\_GC$, mean $\chi^2$, the LDSC intercept with standard error, and the ratio statistic. An LDSC intercept close to 1 was interpreted as evidence that inflation in association statistics was predominantly attributable to polygenicity rather than population stratification or cryptic relatedness.

### 11. Colocalization

Bayesian colocalization was performed using coloc.abf (27) between PA diameter association signals and tissue-specific eQTL datasets relevant to pulmonary vascular and cardiac biology. Uniformly processed GTEx v8 summary statistics (28) were obtained via the eQTL Catalogue (29) and evaluated across six tissues (lung, coronary artery, tibial artery, aorta, heart atrial appendage, and left ventricle), supplemented by lung eQTL data from the Lung Tissue Research Consortium (LTRC) (30). Summary statistics within ±500 kb of each COJO lead variant were harmonized by chromosome, genomic position, and allele coding, with strand-ambiguous variants excluded and effect alleles aligned across datasets.

Loci were classified according to the dominant posterior probability: H4 (shared causal variant), H3 (distinct causal variants), or H0–H2. Evidence of colocalization was defined as posterior probability ≥ 0.8 for hypothesis 4 (PP.H4); for colocalized loci, the variant with the highest SNP-level PP.H4 was used to identify the corresponding eQTL gene (GENCODE v38) (31).

Additional colocalization analyses were performed using GWAS summary statistics for pulse pressure (Million Veteran Program) (32) and pulmonary arterial hypertension (33). Plasma protein QTL associations for lead variants were queried in the UK Biobank Pharma Proteomics Project (34).

**12. Open Targets Genetics annotation and phenotype harmonization**

Open Targets Genetics (35) is an integrative resource that prioritizes likely causal genes at GWAS loci by combining statistical fine-mapping, molecular QTL data, functional genomics annotations, and gene proximity information. The locus-to-gene (L2G) score is a machine-learning metric that integrates multiple genomic features to estimate the likelihood that a gene is the causal effector gene at a given locus. Scores were extracted for all signals, and genes with L2G ≥ 0.5 were considered high-confidence candidates. Regulatory QTL evidence was evaluated across expression QTL (eQTL), protein QTL (pQTL), splicing QTL (sQTL), single-cell expression QTL (sc-eQTL), and transcript usage QTL (tuQTL) datasets, with relationships classified as cis, trans, or mixed.

As Open Targets Genetics aggregates results across thousands of GWAS studies and traits, related phenotypes frequently appeared under multiple study-specific labels. To facilitate cross-trait comparisons and pleiotropy analyses, phenotype annotations were harmonized into biologically coherent categories using a keyword-based pipeline. Trait labels underwent text normalization to remove study-specific identifiers before assignment into one of nine broad phenotype domains: Lung/Respiratory, Cardiovascular, Metabolic, Hematology, Body Composition, Immune/Inflammatory, Brain/Neurological, Cancer, and Other/Multisystem. Harmonized labels were used to summarize pleiotropic associations across organ systems.

**13. Evidence integration and effector-gene prioritization**

To prioritize candidate effector genes for conditionally independent PA diameter signals, we integrated evidence from five complementary analytical layers: (1) a qualifying coding variant within the lead's 95% credible set (missense or nonsense; PIP ≥ 0.10), (2) tissue eQTL colocalization (GTEx v8; PP.H4 ≥ 0.80), (3) Open Targets locus-to-gene score (L2G ≥ 0.50), (4) enhancer-to-gene score (E2G ≥ 0.60), and (5) molecular QTL evidence in any modality (eQTL, pQTL, sQTL, sc-eQTL, tuQTL). Enhancer-to-gene scores from the E2G Consortium (36,37) link regulatory variants to predicted target genes based on chromatin accessibility, histone modification, and three-dimensional chromatin interaction data; scores ≥ 0.6 were retained.

For each signal, the prioritized effector gene was defined by the highest-ranking qualifying evidence source, using the hierarchy coding variant > eQTL colocalization > L2G > E2G > molecular QTL. When no evidence source met prespecified thresholds, the nearest protein-coding gene (GENCODE v38) was assigned. The convergence score was defined as the number of qualifying qualifying evidence sources supporting the prioritized gene, and concordance with the nearest protein-coding gene was recorded for each signal. Colocalization of PA diameter signals with pulse pressure was assessed and reported as a separate analysis but was not incorporated into the effector-gene prioritization framework.

#### 14. Single-cell expression

Cellular context relevant to pulmonary vascular remodeling was evaluated using the LungMAP Human Lung CellRef single-cell RNA-sequencing atlas (38) via the ShinyCell interface (https://app.lungmap.net/app/shinycell-human-lung-cellref). Prioritized effector genes were evaluated for cell-type-specific expression across pulmonary vascular-relevant cell populations, including endothelial cells, vascular smooth muscle cells, pericytes, and fibroblasts.

---

### Supplementary Results

#### 1. Rare-variant gene-based analysis

Gene-based burden testing of pulmonary artery diameter identified GEM (ENSG00000164949, chr8) as the only gene reaching Bonferroni-corrected significance in COPDGene (Figure S3; Table S3). The association was driven by seven ultra-rare predicted loss-of-function and deleterious-missense variants captured in the Set2 singleton mask (Table S3 footnote). The association was robust across strata, remained significant in COPD subjects overall (GOLD 1-4) and in moderate-to-severe disease (GOLD 2-4), indicating it was not confined to a single subgroup. No additional gene reached significance in any analysis, including the combined COPDGene + ECLIPSE cohort (Figure S3). Quantile–quantile plots showed no evidence of genomic inflation (Figure S3). GEM was not significant in the omnibus GENE_P test, consistent with the association being driven primarily by singleton variants and diluted by aggregation across variant masks and tests.

### 2. PA/A ratio and PA Enlargement

For the PA-to-aorta ratio, Stage 1 discovery in the combined COPDGene and ECLIPSE cohort identified a single genome-wide significant locus on chromosome 2 near *ACYP2* (lead variant rs1363065, $P = 2.4\times10^{-8}$; Figure S2). No variant reached genome-wide significance for the binary PA-enlargement trait (PA/A > 1). In the Stage 2 meta-analysis across the cohorts with aortic measurements (COPDGene, ECLIPSE, and the UK Biobank), genome-wide significant PA/A loci are reported in Table S5. Cross-cohort effect-size concordance for PA/A was strong: among variants genome-wide significant in either or both settings (n = 516), effect estimates were highly correlated between the COPD-enriched and UK Biobank cohorts (Pearson r = 0.89, $P = 2.2\times10^{-16}$; Figure S5). Consistent with a partly shared genetic basis, the cross-cohort genetic correlation for PA/A was moderate (rg = 0.57) and significantly lower than the near-unity correlation observed for PA diameter, likely reflecting additional biological and technical heterogeneity introduced by the aortic component of the phenotype.

### 3. Heritability and genetic correlation

Observed-scale SNP heritability in the UK Biobank was substantial for both PA diameter ($h^2 = 0.286$, SE 0.022) and PA/A ($h^2 = 0.207$, SE 0.021). Heritability estimates in the COPD-enriched cohorts were lower and less precise (PA diameter: $h^2 = 0.107$, SE 0.036; PA/A: $h^2 = 0.131$, SE 0.035), likely reflecting smaller sample size and greater environmental and disease-related heterogeneity in smoking-exposed populations. Across all analyses, LD score regression intercepts remained near 1.0, indicating minimal inflation from population stratification or other confounding and suggesting that the observed association signals were driven predominantly by true polygenicity (Table S6).

Genetic correlation analyses demonstrated extensive sharing of genetic determinants between the COPD-enriched and population-based cohorts. For PA diameter, the genetic correlation was approximately 1 (rg = 1.02, SE 0.17, $P = 3.0\times10^{-9}$), indicating highly consistent genetic effects despite marked differences in smoking exposure and COPD burden. For PA/A, the genetic correlation was significant but more moderate (rg = 0.57, SE 0.12, $P = 4.7\times10^{-6}$) and significantly lower than 1 ($P = 5\times10^{-4}$), potentially reflecting additional biological and technical heterogeneity introduced by the aortic component of the phenotype. Together, these findings indicate that pulmonary vascular structure is heritable and largely governed by shared genetic determinants across populations with markedly different COPD burden and environmental exposures.

### 4. Fine-mapping and credible-set annotation

Fine-mapping markedly narrowed the set of plausible causal variants at many loci, with several signals resolving to compact credible sets and three loci (*MECOM*, *ABCC8*, and *PDE3A*) achieving single-variant resolution. Broader credible sets persisted in regions with more complex local linkage disequilibrium structure and smaller effect sizes. The complete molecular-consequence annotation of credible-set variants, including coding and regulatory variants underlying prioritized signals, is provided in Tables S9 and

S10. At the *SH2B3* locus, the same credible set additionally contained a 3′UTR variant at PIP=0.14, suggesting both coding and regulatory mechanisms. Five promoter variants reached PIP≥0.37, including a single-variant credible set at PIP=1.00 in the *ABCC8* region.

### 5. Colocalization

At the locus tagged by the lead variant 8:42467247:T:C, colocalization prioritized *SMIM19* rather than the nearest protein-coding gene, *SLC20A2*, highlighting the limitations of positional annotation alone and underscoring the importance of regulatory evidence for effector-gene prioritization. Representative regional colocalization (LocusCompare) plots are shown in Figure S8.

### 6. Open Targets prioritization

Querying Open Targets Genetics were available for 39 of the 41 lead variants. *ABCC8* emerged as one of the highest-confidence effector genes, supported by Open Targets-native summaries, including L2G score against fine-mapping posterior probability (Figure S10) and multi-modality QTL convergence (Figure S11). Per-signal integrated evidence is provided in Table S12.

### 7. Cellular expression of prioritized effector genes

In the LungMAP Human Lung CellRef single-nucleus atlas, prioritized effector genes showed enriched expression in pulmonary vascular-relevant cell populations, including endothelial cells, vascular smooth muscle cells, pericytes, and fibroblasts (Figure S12), linking the genetic associations to cellular compartments involved in pulmonary vascular remodeling.

### Supplementary Figures (Figures S1–S12)

**Figure S1. Participant flow for the COPDGene and ECLIPSE discovery cohorts.** Flow diagrams for COPDGene (A) and ECLIPSE (B): enrolled participants and sequential exclusions for missing PA measurements, missing model covariates, and genotype-level quality control, ending in the analytic sample for the common-variant association analysis.

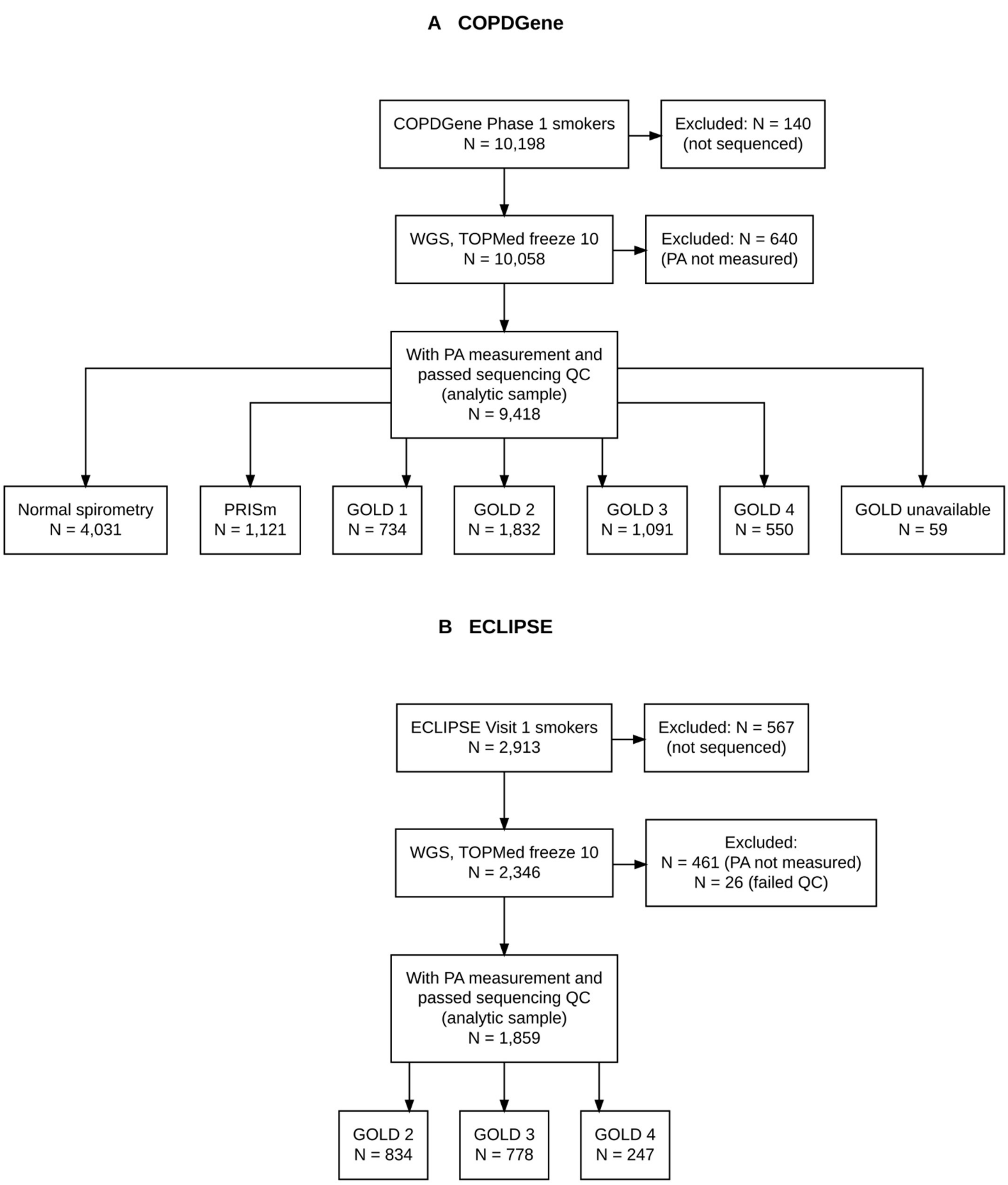
A COPDGene
COPDGene Phase 1 smokers
N = 10,198
Excluded: N = 140
(not sequenced)
WGS, TOPMed freeze 10
N = 10,058
Excluded: N = 640
(PA not measured)
With PA measurement and
passed sequencing QC
(analytic sample)
N = 9,418
Normal spirometry
N = 4,031
PRISm
N = 1,121
GOLD 1
N = 734
GOLD 2
N = 1,832
GOLD 3
N = 1,091
GOLD 4
N = 550
GOLD unavailable
N = 59
B ECLIPSE
ECLIPSE Visit 1 smokers
N = 2,913
Excluded: N = 567
(not sequenced)
WGS, TOPMed freeze 10
N = 2,346
Excluded:
N = 461 (PA not measured)
N = 26 (failed QC)
With PA measurement and
passed sequencing QC
(analytic sample)
N = 1,859
GOLD 2
N = 834
GOLD 3
N = 778
GOLD 4
N = 247

**Figure S2. Stage 1 common-variant association results for PA and PA/A and COPDGene Phase 2 results for PA.** Manhattan and quantile–quantile (QQ) plots for PA diameter (A, B) and the PA/A ratio (C, D) in the combined COPDGene and ECLIPSE discovery cohort (n = 11,277), and for the COPDGene Phase 2 analysis (E, F; n = 4,848). Variants within genome-wide significant loci (±500 kb) are highlighted in red; lead variants are shown as diamonds annotated by the nearest protein-coding gene. The solid line denotes genome-wide significance ($P = 5\times10^{-8}$) and the dashed line the suggestive threshold ($P = 1\times10^{-5}$). For PA/A, a chromosome 2 locus near ACYP2 (rs1363065) reached genome-wide significance; in Phase 2 the ANO1 signal reproduced, with the lead variant in strong linkage disequilibrium ($r^2 = 0.99$) with rs386829462.

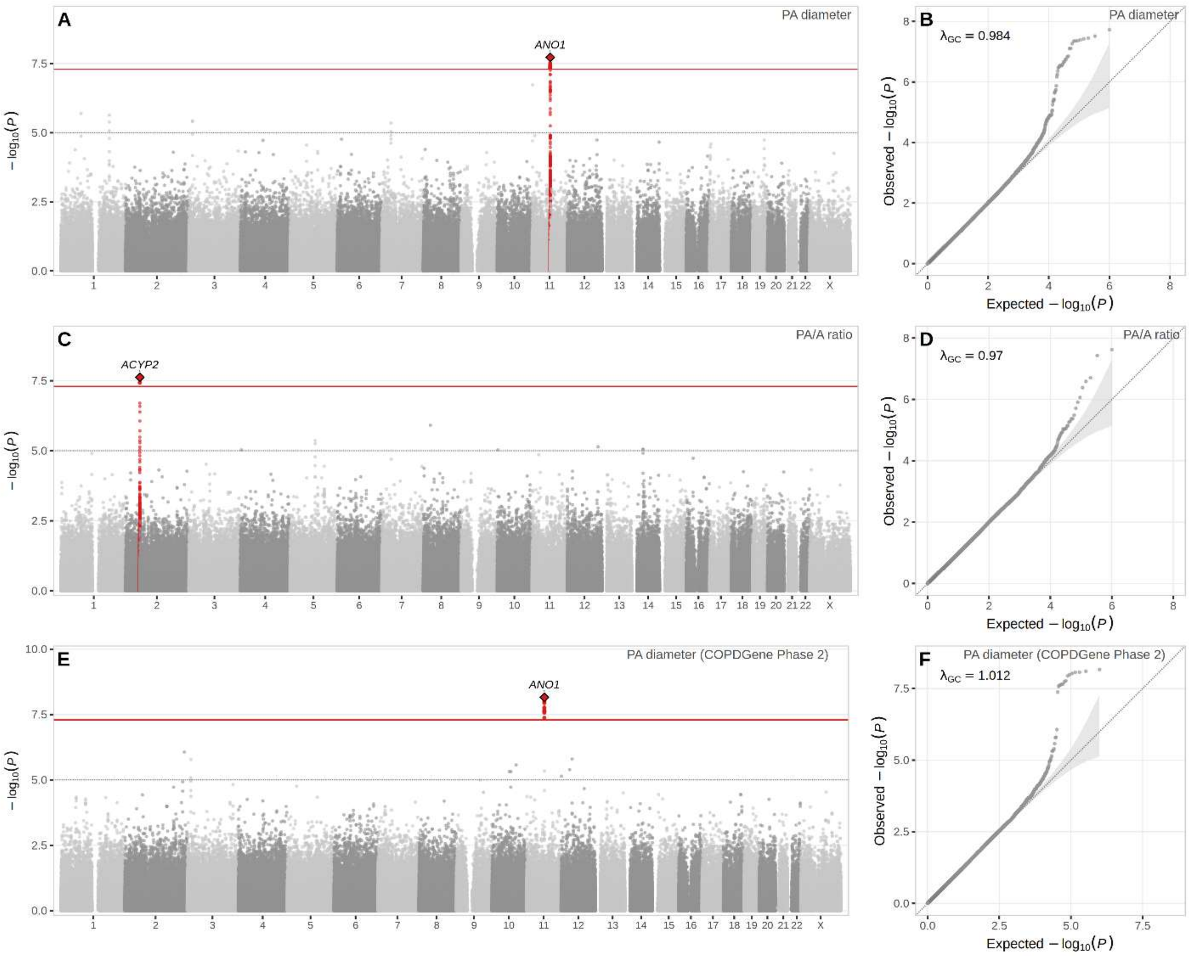
A
PA diameter
ANO1
$-\log_{10}(P)$
B
PA diameter
$\lambda_{GC}$ = 0.984
Observed $-\log_{10}(P)$
Expected $-\log_{10}(P)$
C
PA/A ratio
ACYP2
$-\log_{10}(P)$
D
PA/A ratio
$\lambda_{GC}$ = 0.97
Observed $-\log_{10}(P)$
Expected $-\log_{10}(P)$
E
PA diameter (COPDGene Phase 2)
ANO1
$-\log_{10}(P)$
F
PA diameter (COPDGene Phase 2)
$\lambda_{GC}$ = 1.012
Observed $-\log_{10}(P)$
Expected $-\log_{10}(P)$

**Figure S3. Gene-based rare-variant burden testing of PA diameter.** (A) Manhattan plots: genome-wide $-\log_{10}$(P) from gene-based burden testing of PA diameter for COPDGene, COPDGene COPD GOLD1-4, and COPDGene GOLD2-4. Dashed line, analysis-specific Bonferroni threshold (P < 0.05 / genes tested). (B) Quantile–quantile plots of the gene-based test statistics for the analyses in (A). GEM reached Bonferroni-corrected significance for PA diameter in COPDGene.

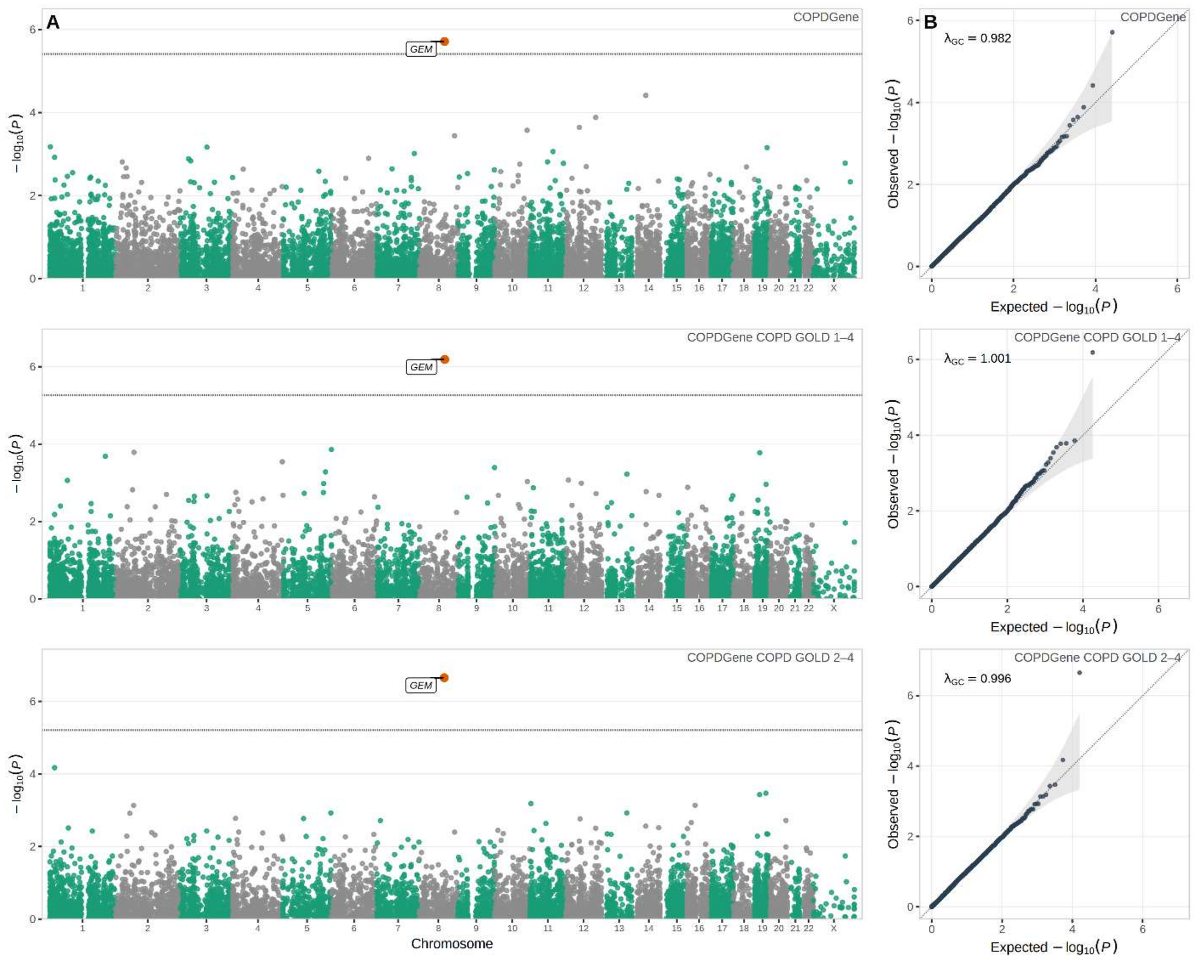

**Figure S4. Stage 2 meta-analysis of PA diameter and the PA/A ratio.** Manhattan and QQ plots for the Stage 2 fixed-effects meta-analysis (COPDGene + ECLIPSE + UK Biobank, n = 48,350) for PA diameter (A, B) and PA/A (C, D). Highlighting, lead-variant annotation, and significance thresholds are as in Figure S2. Thirty-seven genome-wide significant PA diameter loci were identified.

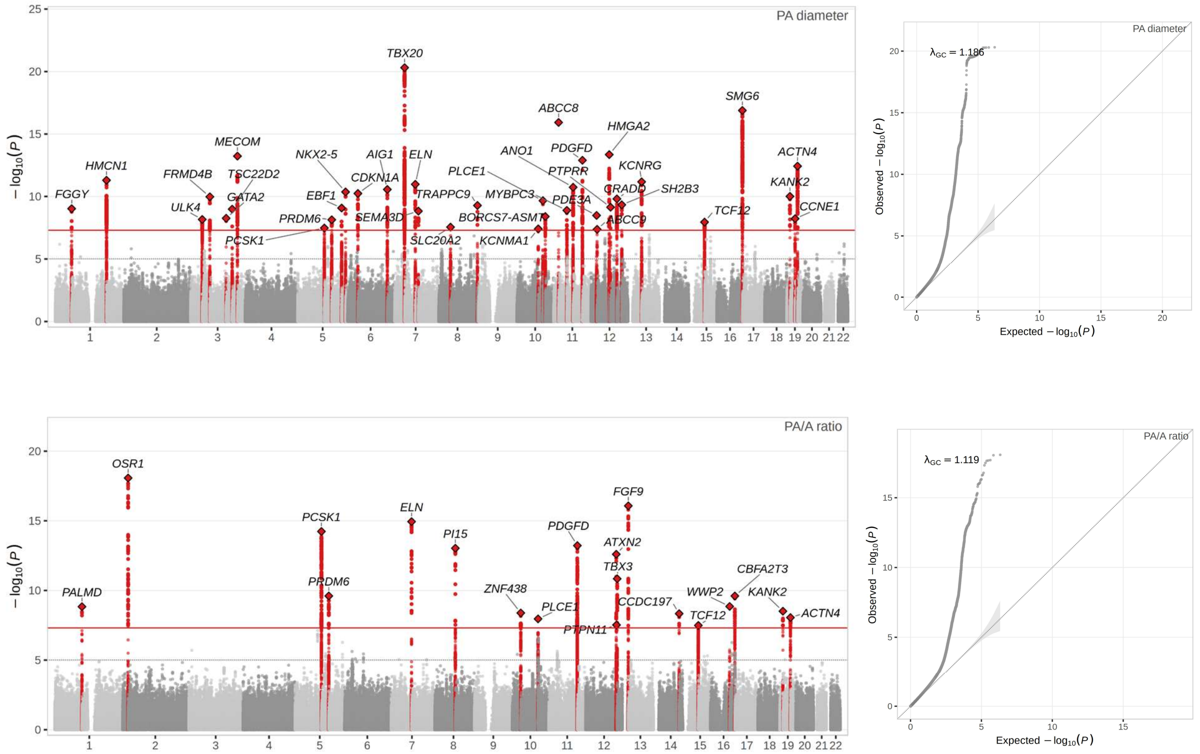

**Figure S5. Cross-cohort effect-size concordance for the PA/A ratio.** Effect estimates (β) for the PA/A ratio compared between the COPD-enriched and UK Biobank settings, for variants genome-wide significant in either or both datasets (n = 516); the diagonal indicates perfect concordance. Effect estimates were strongly correlated (Pearson r = 0.83, P = $2.2\times10^{-16}$).

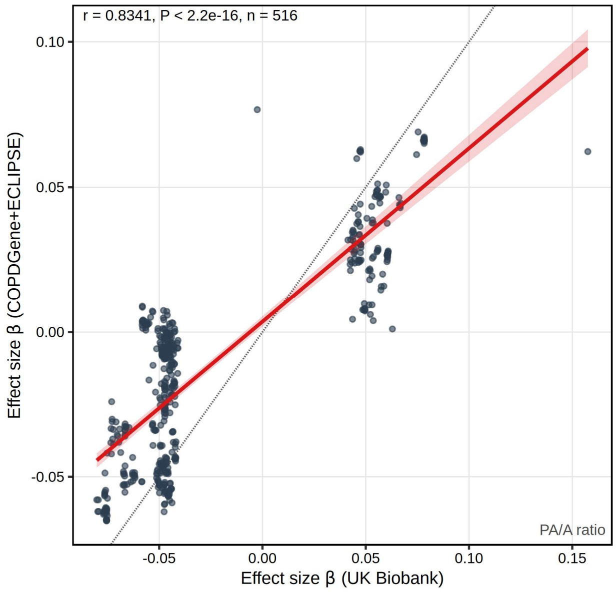

**Figure S6. Four-cohort joint meta-analysis and European-ancestry-restricted analysis.** (A) QQ plot for the four-cohort joint meta-analysis of PA diameter (n = 51,639; the corresponding Manhattan is main-text Figure 3). (B) Manhattan and (C) QQ plot for the analysis restricted to participants of self-reported European ancestry, which served as the basis for conditional and joint analysis and fine-mapping. Highlighting and thresholds are as in Figure S4.

**A.**

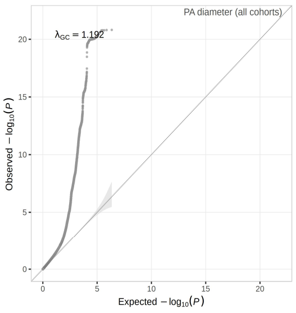


**B.**

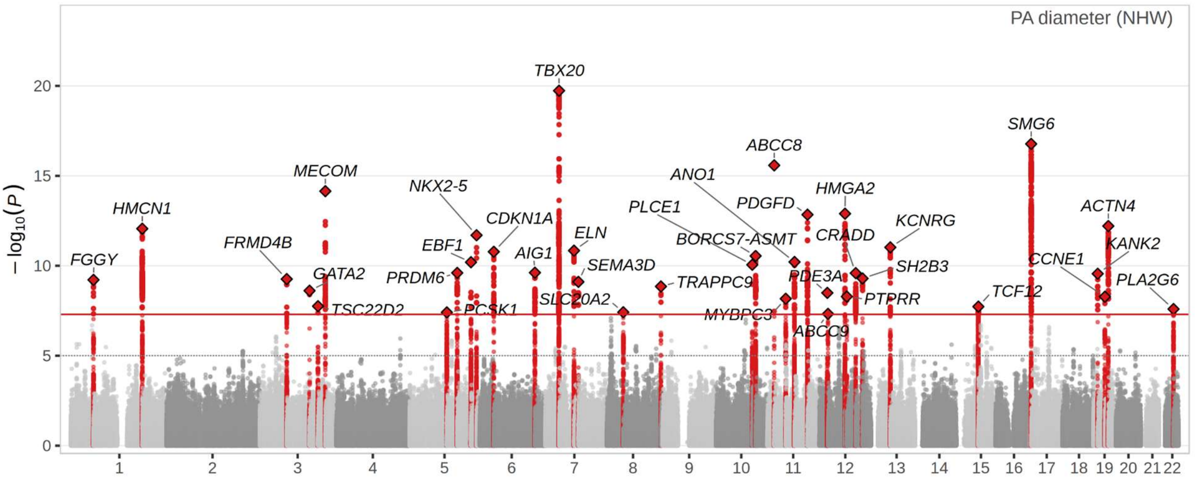

**C.**

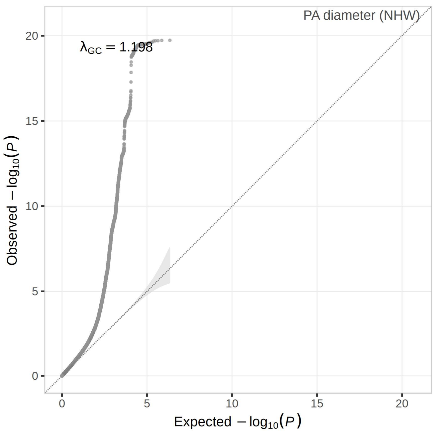


**Figure S7. Colocalization across PA diameter signals.** Heatmap of colocalization posterior probabilities (PP.H4) across PA diameter signals for tissue eQTLs (GTEx v8: lung, coronary artery, tibial artery, aorta, heart atrial appendage, left ventricle, whole blood; and the Lung Tissue Research Consortium and for pulse pressure. Each cell is annotated with its PP.H4; row labels give the signal and lead variant. Colocalization was defined as PP.H4 ≥ 0.8.

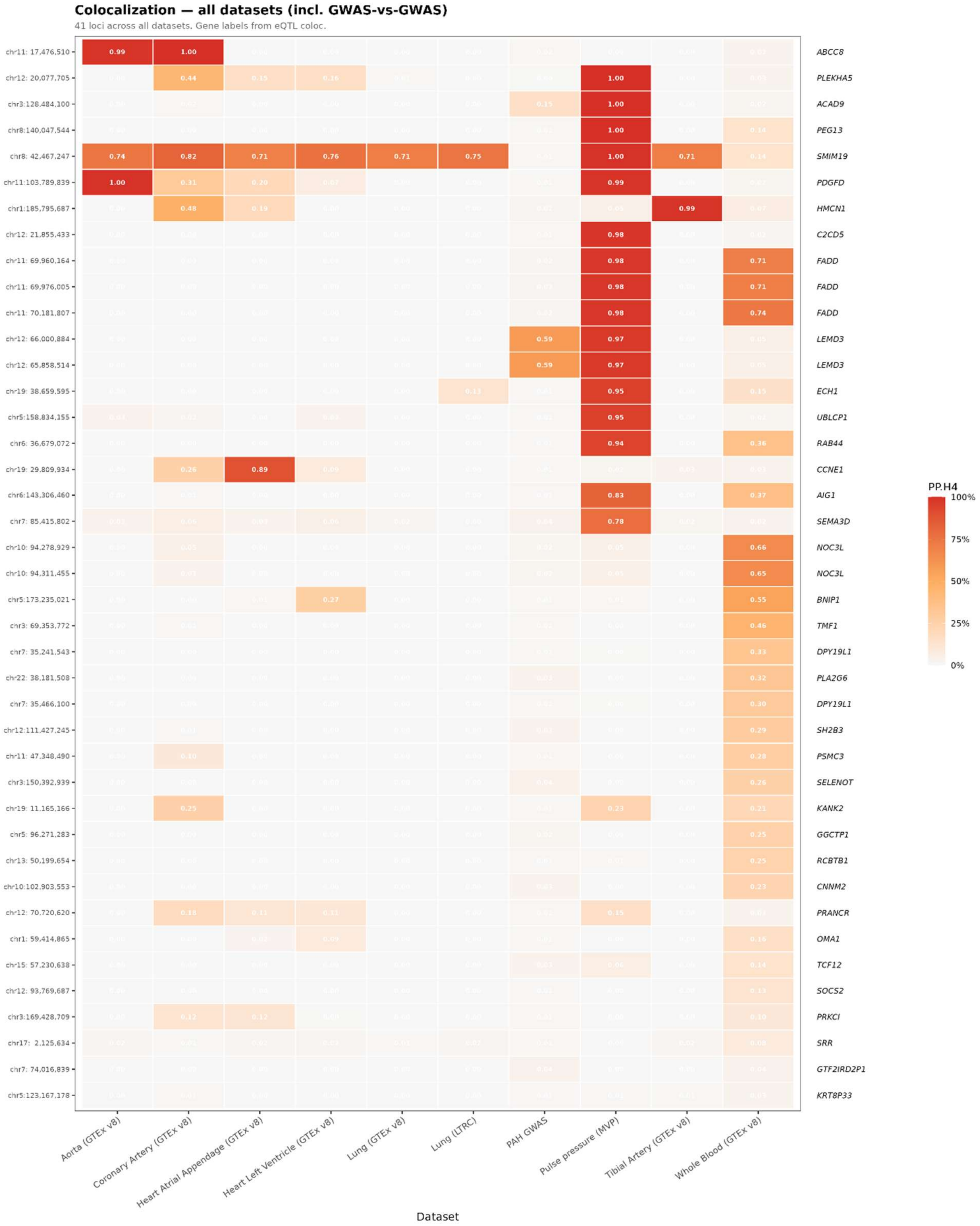
Colocalization — all datasets (incl. GWAS-vs-GWAS)
41 loci across all datasets. Gene labels from eQTL coloc.
chr11: 17,476,510
chr12: 20,077,705
chr3:128,484,100
chr8:140,047,544
chr8: 42,467,247
chr11:103,789,839
chr1:185,795,687
chr12: 21,855,433
chr11: 69,960,164
chr11: 69,976,005
chr11: 70,181,807
chr12: 66,000,884
chr12: 65,858,514
chr19: 38,659,595
chr5:158,834,155
chr6: 36,679,072
chr19: 29,809,934
chr6:143,306,460
chr7: 85,415,802
chr10: 94,278,929
chr10: 94,311,455
chr5:173,235,021
chr3: 69,353,772
chr7: 35,241,543
chr22: 38,181,508
chr7: 35,466,100
chr12:111,427,245
chr11: 47,348,490
chr3:150,392,939
chr19: 11,165,166
chr5: 96,271,283
chr13: 50,199,654
chr10:102,903,553
chr12: 70,720,620
chr1: 59,414,865
chr15: 57,230,638
chr12: 93,769,687
chr3:169,428,709
chr17: 2,125,634
chr7: 74,016,839
chr5:123,167,178
ABCC8
PLEKHA5
ACAD9
PEG13
SMIM19
PDGFD
HMCN1
C2CD5
FADD
FADD
FADD
LEMD3
LEMD3
ECH1
UBLCP1
RAB44
CCNE1
AIG1
SEMA3D
NOC3L
NOC3L
BNIP1
TMF1
DPY19L1
PLA2G6
DPY19L1
SH2B3
PSMC3
SELENOT
KANK2
GGCTP1
RCBTB1
CNNM2
PRANCR
OMA1
TCF12
SOCS2
PRKCI
SRR
GTF2IRD2P1
KRT8P33
PP.H4
100%
75%
50%
25%
0%
Aorta (GTEx v8)
Coronary Artery (GTEx v8)
Heart Atrial Appendage (GTEx v8)
Heart Left Ventricle (GTEx v8)
Lung (GTEx v8)
Lung (LTRC)
PAH GWAS
Pulse pressure (MVP)
Tibial Artery (GTEx v8)
Whole Blood (GTEx v8)
Dataset

**Figure S8. Representative regional colocalization (LocusCompare) plots.** LocusCompare plots at representative colocalized loci (PP.H4 ≥ 0.8) comparing PA diameter association signals and PP GWAS; points are colored by linkage disequilibrium ($r^2$) relative to the candidate shared causal variant.

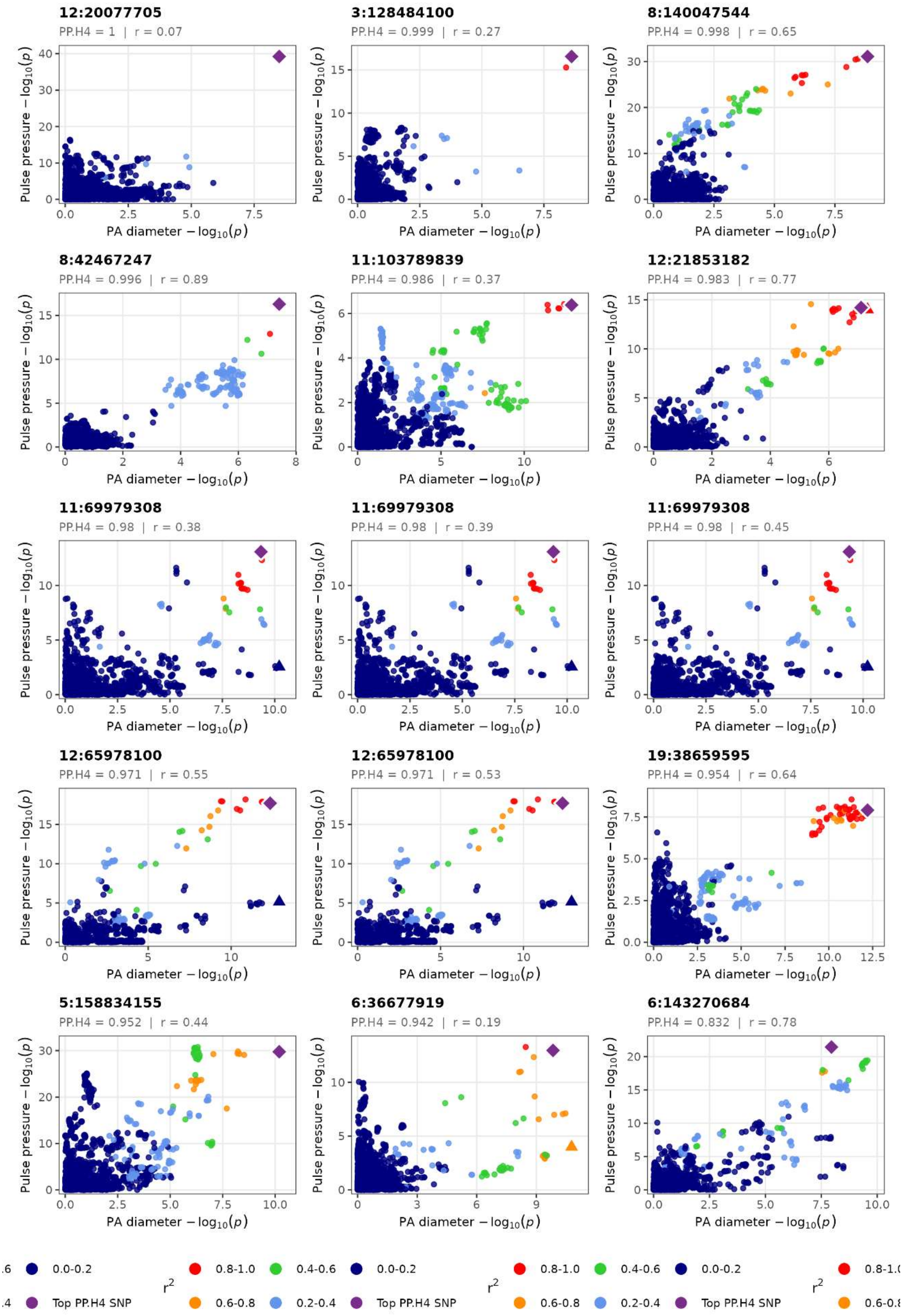

12:20077705
PP.H4 = 1 | r = 0.07
3:128484100
PP.H4 = 0.999 | r = 0.27
8:140047544
PP.H4 = 0.998 | r = 0.65
8:42467247
PP.H4 = 0.996 | r = 0.89
11:103789839
PP.H4 = 0.986 | r = 0.37
12:21853182
PP.H4 = 0.983 | r = 0.77
11:69979308
PP.H4 = 0.98 | r = 0.38
11:69979308
PP.H4 = 0.98 | r = 0.39
11:69979308
PP.H4 = 0.98 | r = 0.45
12:65978100
PP.H4 = 0.971 | r = 0.55
12:65978100
PP.H4 = 0.971 | r = 0.53
19:38659595
PP.H4 = 0.954 | r = 0.64
5:158834155
PP.H4 = 0.952 | r = 0.44
6:36677919
PP.H4 = 0.942 | r = 0.19
6:143270684
PP.H4 = 0.832 | r = 0.78
Pulse pressure $-\log_{10}(p)$
PA diameter $-\log_{10}(p)$
$r^2$
0.8-1.0
0.6-0.8
0.4-0.6
0.2-0.4
0.0-0.2
Top PP.H4 SNP

**Figure S9. Regional association (LocusZoom) plots for the remaining genomic risk loci.** LocusZoom plots for the genomic risk loci not shown in main-text Figure 7. Each panel shows $-\log_{10}(P)$ for PA diameter against genomic position, points colored by linkage disequilibrium ($r^2$) with the lead variant, gene models below, and recombination rate on the right axis. Linkage disequilibrium was estimated from the in-sample self-reported European-ancestry reference panel (COPDGene and ECLIPSE).

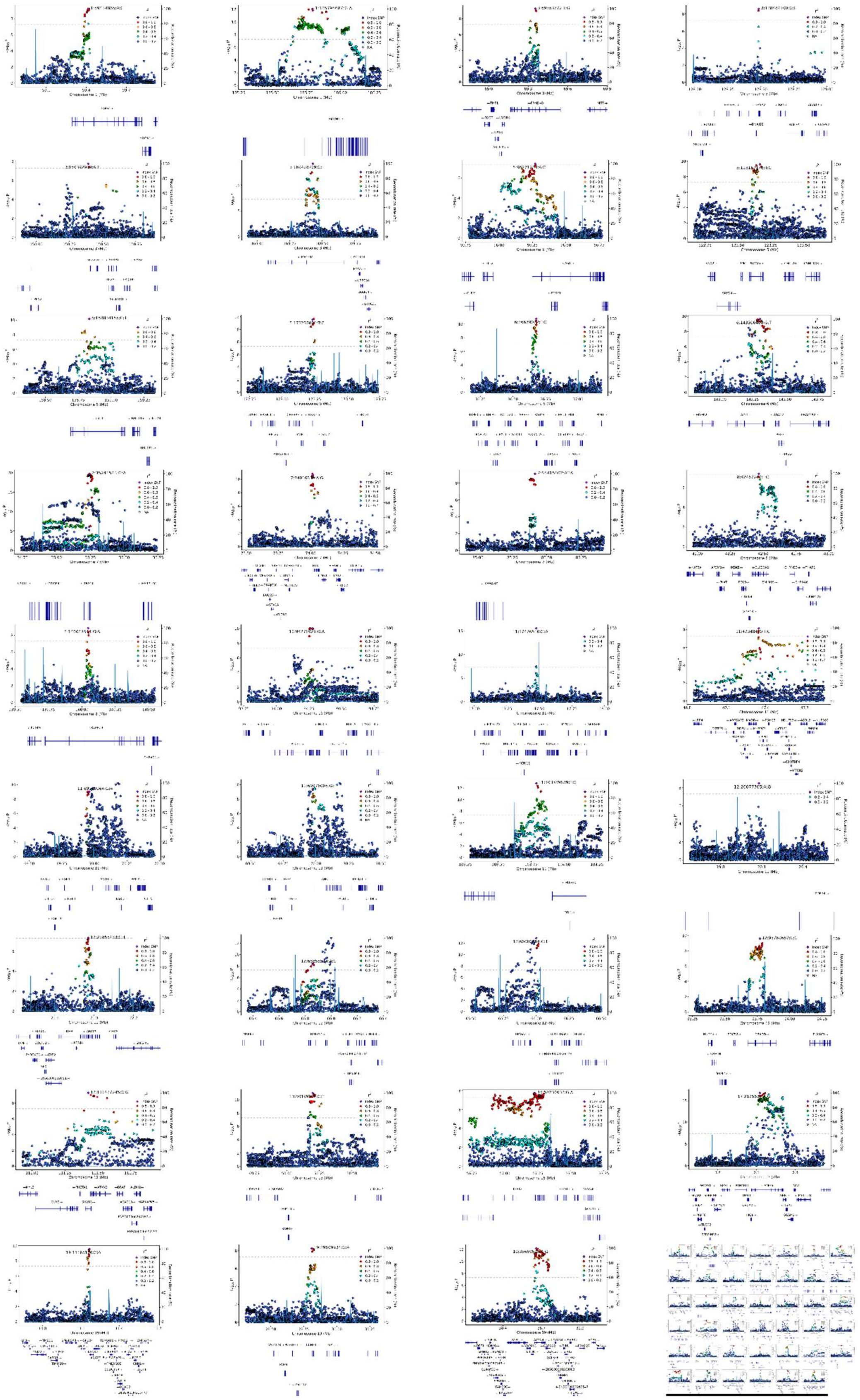

**Figure S10. Open Targets Genetics locus-to-gene prioritization.** Bubble plot of candidate effector genes prioritized through Open Targets Genetics. The x-axis shows the maximum locus-to-gene (L2G) score and the y-axis the maximum fine-mapping posterior probability; bubble size encodes the number of associated cardiovascular traits and color the primary cardiovascular subcategory. *ABCC8* was among the highest-confidence genes.

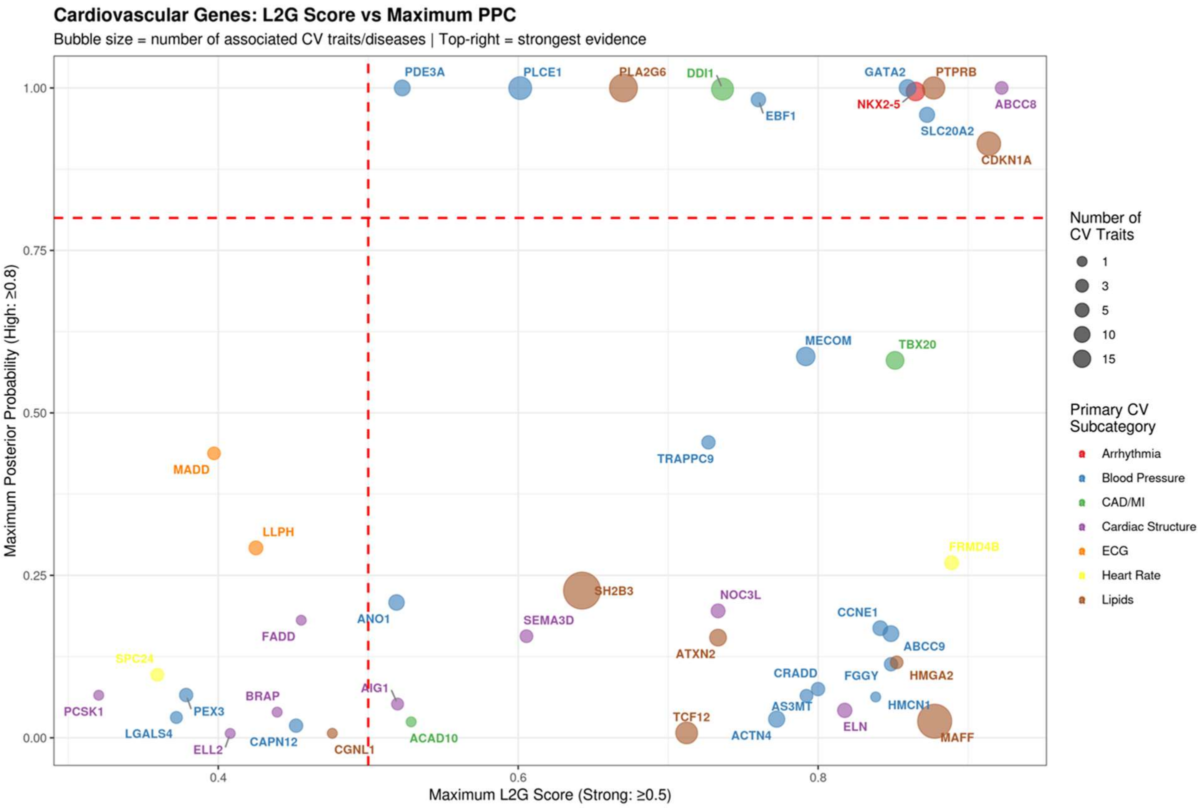

**Figure S11. Convergent QTL evidence supporting candidate effector genes.** Alluvial plot of convergent candidate effector-gene nominations from Open Targets Genetics. Each stream represents an independent association signal and tracks its supporting QTL evidence (eQTL, pQTL, sQTL, sc-eQTL, tuQTL), prioritized effector gene, and associated organ or disease category.

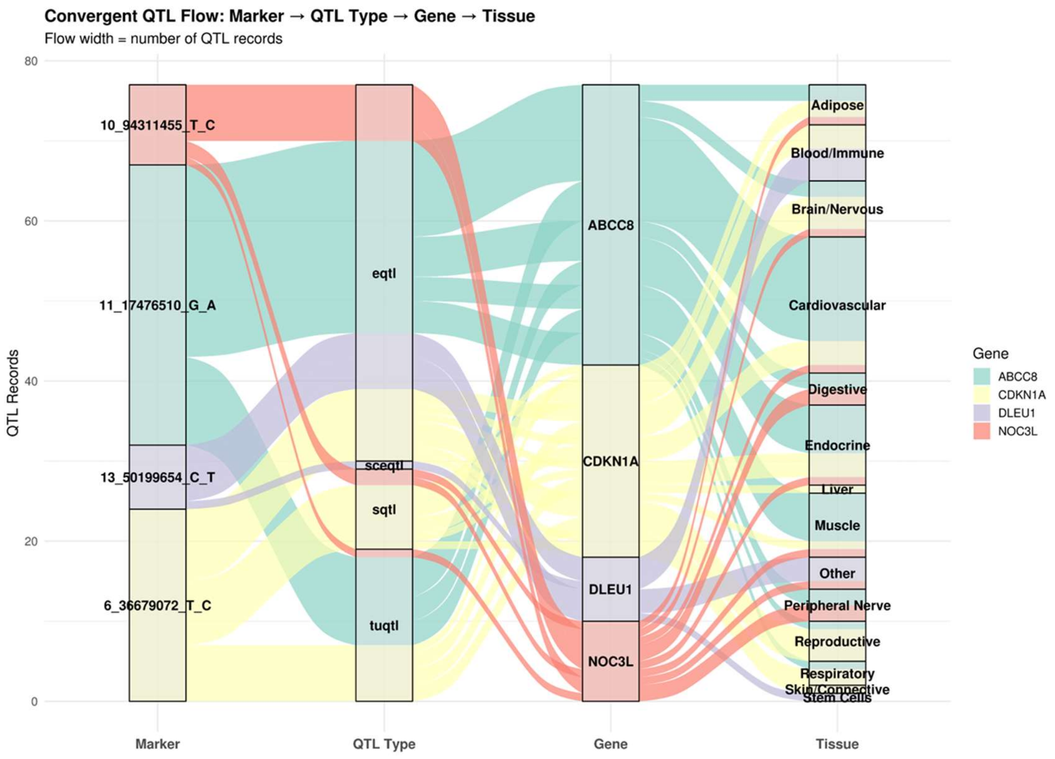

**Figure S12. Cell-type-specific expression of prioritized effector genes.** Dot plot of prioritized effector-gene expression across pulmonary vascular-relevant cell populations in the LungMAP Human Lung CellRef single-nucleus atlas: mean expression (color) and proportion of expressing cells (dot size) across endothelial cells, vascular smooth muscle cells, pericytes, and fibroblasts.

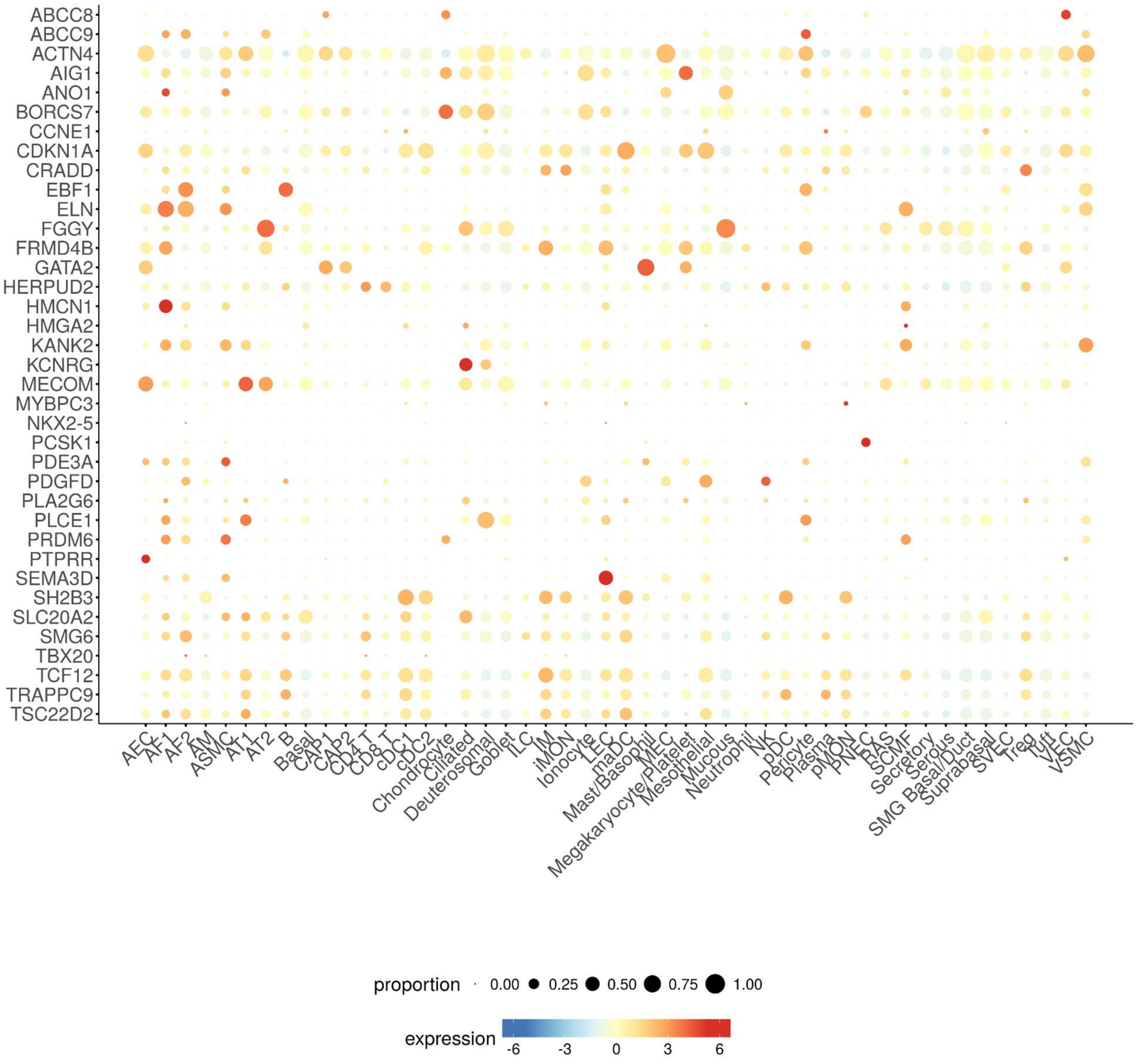

### Supplementary Table Legends (Tables S1–S13)

**Table S1. Additional characteristics of the COPDGene and ECLIPSE cohorts.** Additional demographic, anthropometric, spirometric, and PA imaging characteristics for COPDGene and ECLIPSE, reported overall for each cohort. Continuous variables are presented as median (interquartile range) and categorical variables as n (%). Four-cohort baseline characteristics are given in main-text Table 1.

**Table S2. COPDGene Phase 2 characteristics.** Characteristics of the COPDGene Phase 2 participants by sex. Continuous variables, median (interquartile range); categorical variables, n (%).

**Table S3. Suggestive gene-based rare-variant associations.** Gene-based rare-variant burden results at suggestive significance for PA diameter listing the exome-wide significant associations of GEM with PA diameter in COPDGene.

**Table S4. Genome-wide significant PA diameter loci, Stage 2 meta-analysis.** The 37 genome-wide significant PA diameter loci from the Stage 2 meta-analysis (COPDGene + ECLIPSE + UK Biobank), with lead variant, chromosome and position (GRCh38), effect and other alleles, effect-allele frequency, effect estimate, P value, nearest protein-coding gene, and known/novel status.

**Table S5. Genome-wide significant loci for the PA/A ratio, Stage 2 meta-analysis.** Genome-wide significant PA/A loci from the Stage 2 meta-analysis (COPDGene + ECLIPSE + UK Biobank), with lead variant, chromosome and position (GRCh38), effect and other alleles, effect-allele frequency, effect estimate, P value, nearest protein-coding gene, and known/novel status.

**Table S6. SNP-based heritability and cross-cohort genetic correlation (LD score regression).** Observed-scale SNP heritability for PA diameter and the PA/A ratio in the UK Biobank and COPD-enriched (COPDGene + ECLIPSE) cohorts, with λ_GC, mean $\chi^2$, LDSC intercept, and ratio statistic, and genetic correlations (rg) between the cohorts.

**Table S7. Replication of Stage 2 lead variants in the Framingham Heart Study.** Replication of Stage 2 PA diameter lead variants in the Framingham Heart Study: lead variant, effect and other alleles, the discovery effect estimate, and the FHS effect estimate, standard error, P value, and directional concordance.

**Table S8. Conditionally independent PA diameter signals (GCTA-COJO).** The 41 conditionally independent signals identified by GCTA-COJO (LD $r^2 < 0.9$) across the 36 self-reported European-ancestry loci, with locus, lead variant, position (GRCh38), alleles, effect-allele frequency, marginal and conditional effect estimates and P values, and the nearest protein-coding gene.

**Table S9. SuSiE-RSS fine-mapping credible sets.** The 45 95% credible sets across the 36 loci: locus, signal, credible-set size, lead variant, posterior inclusion probability, and credible-set purity.

**Table S10. Predicted molecular consequence of credible-set variants.** Molecular-consequence annotation (VariantAnnotation, GRCh38) of variants in 95% credible sets, each assigned a single tier using the hierarchy nonsense > missense > splice site > 5′UTR > 3′UTR > synonymous > promoter > intron > intergenic.

**Table S11. Open Targets phenotype harmonization.** Mapping of study-specific trait labels to the nine harmonized phenotype domains used to summarize pleiotropic associations.

**Table S12. Integrated effector-gene evidence.** Per-signal integrated effector-gene prioritization across the five evidence layers (coding variant in credible set, eQTL colocalization, L2G, E2G, molecular QTL), with the prioritized effector gene, number of converging layers, and nearest protein-coding gene.

**Table S13. Plasma protein QTL associations (UK Biobank Pharma Proteomics Project).** Plasma protein QTL associations for PA diameter lead variants. The *SH2B3* lead variant rs7310615 was the only variant associated with circulating protein levels, with numerous trans-pQTL associations (including IL-15, EMCN, HGF, OSM, and TGFα).

---
